# Integrated CRISPR-based detection of HIV, tuberculosis, malaria, and resistance-associated variants

**DOI:** 10.64898/2026.09.26.26364094

**Authors:** Ashvi S. Jain, Austin Si, Joshua Yap, Lilith A Schwartz, Cara Susilo, Arumugam Balamurugan, Arrmund Neal, Karine G. Le Roch, Otto Yang, Omai Garner, Mireille Kamariza

## Abstract

HIV, tuberculosis (TB) and malaria are major infectious diseases that frequently co-circulate in high-burden settings, yet their diagnostic pathways remain largely disease-specific, complicating detection of coinfection and integration of antimicrobial-resistance testing. Here we developed HTM CARMEN, a highly multiplexed CRISPR-Cas13a molecular-testing framework that integrates detection of HIV-1, HIV-2, four *Plasmodium* species and *Mycobacterium tuberculosis* (Mtb) with detection of isoniazid-resistance-associated *inhA* and *katG* variants. We find that HTM CARMEN retained target selectivity and single-nucleotide discrimination across *in vitro* experiments and clinically relevant sample matrices, including whole blood, serum, sputum and dried blood spots. Across 20 contrived samples for each condition, the HTM panel supported detection of low-abundance targets across multiple matrices and frequently detected greater number of low-input samples as positive than qPCR under the conditions tested. In contrived *P. falciparum*/HIV-1 and Mtb/HIV-1 coinfections, both pathogens were simultaneously detected across a range of relative target concentrations. Using confirmed-positive clinical samples, HTM CARMEN achieved positive percent agreement of 95% for malaria (19/20), 95% for HIV-1 (19/20) and 100% for Mtb (30/30), with 100% negative percent agreement for each target in the confirmed-negative samples. Both HIV-1 and Mtb were additionally detected in all nine confirmed-positive HIV-TB coinfection specimens. These findings establish the feasibility of integrating pathogen detection, species differentiation, resistance-associated variant interrogation and coinfection detection within a programmable molecular-testing platform, providing a framework for high-throughput integrated testing in co-endemic settings.

## Introduction

Tuberculosis (TB), human immunodeficiency virus (HIV) and malaria collectively account for close to 270-280 million incident cases each year, disproportionately affecting low- and middle-income countries^1–3^. Given this high annual case volume, clinical diagnostic pipelines need to operate efficiently and at high throughput to meet testing demand for rapid disease confirmation and appropriate treatment. However, even where rapid molecular tests are increasingly available, diagnostic and surveillance pathways often remain organized around disease-specific single-pathogen assays and reports, such that each disease is tested and reported independently, potentially causing delays in rapid detection of coinfections^4,5^. Indeed, in many endemic settings, these diseases co-circulate within the same population, and coinfections often affect clinical presentation, complicate treatment decisions, and worsen outcomes^5,6^. The resulting diagnostic uncertainty creates a critical blind spot for integrated case management and for scalable surveillance of coinfections, particularly when symptoms overlap or sample types differ by pathogen and resistance profiles.

Among the best-characterized syndemics is HIV-TB coinfection^7^. HIV increases susceptibility to TB, caused by *Mycobacterium tuberculosis* (Mtb), and is associated with smear-negative and extrapulmonary disease, limiting the utility of sputum-based diagnostics in settings where patients may be unable to produce sputum and culture-based methods are too slow to guide timely treatment decisions^8,9^. In addition, HIV increases the risk of disseminated and extrapulmonary TB, creating opportunities for sensitive blood-based detection approaches in addition to conventional sputum testing^10^.

Malaria further intersects with both HIV and TB in co-endemic populations. Malaria is caused by protozoan parasites of the *Plasmodium* genus, of which *Plasmodium falciparum, Plasmodium vivax, Plasmodium ovale*, and *Plasmodium malariae* represent the four historically predominant human malaria species^3^. These species are responsible for the majority of global malaria burden and differ in geographic distribution, disease severity, relapse potential, and treatment considerations^3,11^. Clinical and epidemiological studies indicate that coinfection can be associated with more severe malaria and that malaria episodes can increase HIV viral load, with implications for onward transmission and patient outcomes^12,13^. TB-malaria coinfection is also clinically important but comparatively understudied. Experimental and modeling work suggests immunological interactions that can increase TB burden during *Plasmodium* coinfection, and disease management is complicated by drug-drug interactions, including reduced efficacy of artemisinin-based combination therapies during rifampicin-containing TB treatment^14^. In the absence of integrated diagnostic tools that can monitor coinfections over the course of therapy, clinicians may miss recurrent or persistent disease or may be forced to make treatment decisions with incomplete microbiological information.

A parallel threat is the continued emergence and spread of antimicrobial resistance. For TB, molecular diagnostic workflows frequently prioritize rifampicin (RIF) resistance, whereas isoniazid (INH) resistance can occur in rifampicin-susceptible disease and may precede multidrug-resistant TB^15,16^. Because INH resistance can be present even when a sample is RIF-susceptible, these RIF-centric diagnostic workflows create a diagnostic gap that can lead to inappropriate treatment selection and delayed recognition of INH resistance.

Together, these challenges underscore the need for diagnostic platforms that can simultaneously capture pathogen identity, coinfection, and antimicrobial resistance within a single, scalable assay. Molecular assays offer high-throughput and scalable capabilities that enable movement beyond the one-test–one-pathogen paradigm. In particular, CRISPR-based diagnostics provide a programmable framework for sequence-specific nucleic-acid detection. Cas13-based systems, such as Specific High Sensitivity Enzymatic Reporter UnLOCKing (SHERLOCK), couple CRISPR RNA (crRNA)-directed target recognition to collateral reporter cleavage, enabling sensitive detection and discrimination of closely related sequences^17–21^. This framework enabled the development of Combinatorial Arrayed Reactions for Multiplexed Evaluation of Nucleic acids (CARMEN), which miniaturizes Cas13-based detection reactions to achieve highly multiplexed nucleic acid detection by combinatorially testing 24 assays against up to 192 samples, supporting up to 4,608 reactions within a single integrated fluidic circuit^22,23^. Previous CARMEN implementations have demonstrated broad viral detection, viral subtyping and drug-resistance mutation discrimination, respiratory virus and variant detection, bacterial identification and antimicrobial resistance gene detection, and multiplex testing of febrile infections^23–25^. However, despite these advances, such technologies have not yet been fully leveraged to address the combined challenges of HIV, TB, and malaria (HTM) coinfection and resistance detection.

Here, we present HTM CARMEN, a multiplex CRISPR-Cas13a panel designed to integrate HIV, TB and malaria detection with *Plasmodium* species differentiation and interrogation of two common INH-resistance-associated Mtb variants within a common molecular-testing workflow. The panel combines newly developed *Plasmodium* and Mtb assays with previously validated HIV-1 and HIV-2 assays^25^, enabling simultaneous interrogation of viral, bacterial and parasitic targets. We evaluated analytical specificity and single-nucleotide discrimination *in vitro*, tested performance across sputum, serum, whole blood and dried blood spots, and compared detection with quantitative PCR (qPCR) across contrived single-pathogen and coinfection samples. Finally, we evaluated HTM CARMEN in clinical specimens confirmed-positive for malaria, HIV or TB, and in clinical specimens with HIV-TB coinfection. Together, these experiments establish the feasibility of integrating pathogen detection, species resolution, resistance-associated variant interrogation and coinfection detection within a programmable high-throughput molecular testing framework.

## Materials and Methods

### Panel Design and Development

Oligonucleotide primers and five crRNA guides were designed to detect conserved regions in all the organism genomes. Briefly, these conserved regions were from the following 5 pathogens: (1) *Plasmodium falciparum* 3D7, (2) *Plasmodium vivax*, (3) *Plasmodium ovale*, (4) *Plasmodium malariae*, (5) *Mycobacterium tuberculosis*. Two pairs of crRNA guides were designed to detect the isoniazid resistance related mutations *inhA* −777 C>T (also referred to as *fabG1* −15 C>T) and *katG* S315T/N in *Mycobacterium tuberculosis*. We leveraged HIV and RNase P assays that have been previously described^25^. For other targets, we used Activity-informed Design with All-inclusive Patrolling of Targets (ADAPT; https://adapt.run/) to design crRNAs^26^. The *P. malariae* 18s ribosomal RNA sequence was constructed from a consensus alignment of various short read accessions of the gene on the National Center for Biotechnology Information’s (NCBI) Nucleotide Database. For the remaining designs, complete gene sequences were collected from the NCBI Nucleotide Database, aligned to identify conserved regions, and passed to ADAPT to produce candidate crRNA designs with >95% coverage. This design combined with the SHERLOCK screening methodology described previously was successful for all targets except *P*.*malariae*. For this organism, we chose to identify regions of its 18s sequence that were unique to *P. malariae* relative to the other *Plasmodium* organisms and iteratively screen various guides that tiled this region until a *P. malariae* specific crRNA was found using SHERLOCK screening. Furthermore, the crRNA designs for *M. tuberculosis* SNP detection (*inhA* and *katG* mutations) were designed using the Cas13a guide design software Building Artificial Diagnostic Guides by Exploring Regions of Sequences (BADGERS; https://github.com/broadinstitute/badgers-cas13)^27^ and subsequently screened for contrasting on and off-target activity. While most crRNA pairs are designed to discern only a single mutation, the *katG* S315T/N crRNA can detect both mutations. It was designed to take advantage of reduced Cas13a activity when a guanine, the wild type allele in the case of *katG* S315, is present at the nucleotide position immediately 5’ to the target^27^. For primer design, highly conserved regions were selected, and NCBI’s Primer-BLAST was used to design primers with a melting temperature of 60°C with default salt concentrations. As a positive control, a previously designed pair of Human RNase P primers and a corresponding crRNA were used. All sequences used in this study can be found in Supplementary Tables S2-S4.

### In vitro transcription of crRNAs

crRNAs were generated by *in vitro* transcription (IVT) as described in Supplementary Methods M2. Guide templates and a T7 promoter primer were ordered as ssDNA oligos from Thermo Fisher Scientific.

### Chemically Synthesized Nucleic Acids

Synthetic double stranded DNA (dsDNA) fragments (gBlocks) corresponding to each target amplicon were ordered from Integrated DNA Technologies. Each gBlock was resuspended in nuclease-free water to a final stock concentration of 10^10^ copies µL^−1^. gBlocks were diluted and used for the screening of crRNAs to determine the best performing designs. Selective gBlocks were further used in CARMEN assays for *in vitro* and contrived sample testing.

### Quantitative Synthetic RNA and DNA

Quantitative synthetic DNA for *P. vivax* and *P. malariae* were purchased from ATCC (PRA-3004SD and PRA-3001SD, respectively). The synthetic DNA samples contained the 18S rRNA gene as well as additional genes from the respective organisms. Quantitative Synthetic RNA for HIV-1 and HIV-2 were also purchased from ATCC (VR-3351SD and VR-3266SD, respectively). A diagnostic plasmid containing the 18s rRNA gene from *P. ovale* was obtained through BEI Resources. (NIAID, NIH: Diagnostic Plasmid Containing the Small Subunit Ribosomal RNA Gene (18S) from *Plasmodium ovale*, MRA-180, contributed by Peter A. Zimmerman). These materials were used as genomic targets for sensitivity and specificity testing in CARMEN assays.

### Genomic DNA Samples

Genomic DNA from *B. microti* and *P. falciparum* were purchased from ATCC (PRA-398DQ and PRA-405D, respectively). Genomic DNA from *T. gondii* was received from the Bradley Research Lab at UCLA.

### Reverse Transcription and PCR Amplification of Nucleic Acids

Reverse primers were ordered from ThermoFisher Scientific and resuspended in nuclease-free water to final concentrations of 100 µM. RNA targets were reverse transcribed and amplified using SuperScript IV Reverse Transcriptase. For the reverse transcription, 10 pg–5 µg total RNA extracted from the sample were mixed with 1 µL of 2 µM reverse primer pool, 1 µL of 10 mM dNTP mix, and nuclease-free water to a volume of 13 µL. This mixture was heated for 5 minutes at 65°C and immediately incubated on ice for 1-2 minutes. This mixture was further combined with 4 µL of 5x SuperScript IV Buffer, 1 µL of 100 mM DTT, 1 µL of RNase Inhibitor, and 1 µL of SuperScript IV Reverse Transcriptase (200 U µL^−1^) and incubated at 55°C for 15 minutes. The resulting complementary DNA was used as a target input in multiplexed PCR.

Primer pools for multiplexed PCR were prepared according to Supplementary Method M3. The primer pools contained 8 primer pairs that encompassed all targets on the HTM panel. Multiplex PCR Master Mix from New England Biolabs was used. Reactions were prepared by mixing 10 µL of 5x Multiplex PCR Master mix, 5 µL of Forward Primer Pool, 5 µL of Reverse Primer Pool, 4 µL of the target input (DNA material or RT product), and nuclease-free water for a total reaction volume of 50 µL. PCR proceeded with a 2-minute initial denaturation at 95°C followed by 40 cycles of 30 seconds at 95°C, 60 seconds at 58°C, and 60 seconds at 72°C. Lastly, there was a final extension step of 5 minutes at 72°C before cooling to 10°C.

### Cas13a Detection via 384-well Fluorescent Plate Reader (SHERLOCK)

Cas13a detection assays for crRNA selection were run using the SHERLOCK methodology^17^. Briefly, gBlocks were diluted to concentrations ranging from 10^4^ to 10^0^ copies µL^−1^ and subsequently PCR amplified using the PCR protocol mentioned in Reverse Transcription and PCR Amplification of Nucleic Acids. For the SHERLOCK detection, a master mix with the following components was prepared: 40 mM Tris-HCl (Thermo Scientific), 1 mM DTT (Thermo Scientific), 1 mM rNTPs (each nucleotide, New England Biolabs), 1 U µL^−1^ RNase inhibitor (New England Biolabs), 22.5 nM *Leptotrichia wadei* (Lwa)Cas13a (GenScript), 1 U µL^−1^ T7 RNA Polymerase (Lucigen), 62.5 nM quenched fluorescent RNA probe (FAM-rUrUrUrUrUrU-3IABkFQ from Integrated DNA Technologies), and 14 mM MgOAc (Invitrogen). This master mix was split into 58.5 µL aliquots in a 96 well PCR plate. 1.67 µL of crRNAs at a concentration of 24 ng µL^−1^ were added to their respective wells followed by 3.15 µL of amplified target. No Target Controls (NTCs) for each crRNA were prepared by adding 3.15 µL of negative PCR control (Amplification control) to the assay. The plate was sealed, vortexed, and briefly centrifuged before transferring to a 384-well plate in 20 µL triplicates. The plate was incubated at 37°C for 3 hours, with fluorescence measurements collected at 5-minute intervals using a QuantStudio 5 Real-Time PCR System. Data was processed using Design and Analysis Software v2.7.0 (Applied Biosystems). crRNA candidates exhibiting high on-target activity and low off-target activity were selected for further validation using CARMEN.

### CARMEN Assays using Standard BioTools Biomark HD

CARMEN assays were performed as previously described using the Biomark HD from Standard BioTools^23,25^. Detection experiments were conducted in 24 assays by 192 sample IFCs (Integrated Fluidic Circuits) combinatorially mixed via the integrated microfluidics channels in each chip. Detection reactions were the result of a combinatorial mix of an assay mix and a sample mix.

The assay mix was prepared at a 1.33X concentration and contained LwaCas13a (Genscript, 600 nM), T7 RNA Polymerase (Lucigen, 1.67 U uL^−1^), Assay Loading Buffer (Standard Biotools, 1.33X) and nuclease-free water. It was split into 24 PCR tubes and prepared crRNAs were added to their respective assay mixes in a ratio of 1:3 to a final concentration of 450 nM. The resulting assay mixes were loaded into the assay wells in the IFCs.

The sample mix was prepared at a 1.5X concentration and contained T7 Buffer (Lucigen, 1.5X), a custom sample buffer (66.66 mM Tris-HCl, 8.33 mM NaCl, 15 mM MgCl_2_, 1.67 mM DTT, 1.67w/v% PEG-8K, nuclease-free water), 1.5 U µL^−1^ RNase Inhibitor (NEB), 1.5 mM rNTPs (NEB), 0.75 mM quenched fluorescent RNA probe (FAM-rUrUrUrUrUrU-3IABkFQ from Integrated DNA Technologies), 1.5X ROX reference dye (Invitrogen), and 1.5X GE Buffer (Standard BioTools). It was split across two 96-well plates and amplified samples were added to the respective wells in a 1:2 ratio with the sample mix. The resulting sample mixes were loaded into both sides of the IFC, comprising a total of 192 sample wells.

The 192.24 IFC was loaded onto the IFC Controller RX (Standard BioTools), where the assay and sample mixes were combinatorially mixed according to the manufacturer’s instructions. Following priming and mixing, the IFC was transferred to the Biomark HD, and fluorescence images were collected at 5-minute intervals for 3 hours at 37°C.

### *In vitro* Cas13a Detection Sensitivity on Biomark HD

Genomic material corresponding to each target was diluted in nuclease-free water to final concentrations of 10^4^, 10^3^, 10^2^, 10^1^, 10^0^, and 10^−1^ copies µL^−1^ and tested using the Biomark HD. Samples were tested in technical duplicate. Reaction kinetics were compared with the corresponding crRNA-specific diagnostic thresholds to determine the lowest concentration detected by each crRNA. Determination of the diagnostic positivity threshold is described in the Data Analysis and Diagnostic Call Criteria section.

### Nucleic Acid Extraction Methods

Genetic material was extracted from *M. tuberculosis, P. falciparum*, and HIV-1 according to the following methods.

#### *M. tuberculosis* gDNA Extraction

*M. tuberculosis* gDNA was extracted via a bead beating step prior to use of the QIAamp DNA Mini Kit. For each preparation, 10 mL of log phase *M. tuberculosis* culture was pelleted (10 minutes at 5000 x g) and isolated from its supernatant. Isolated cells were resuspended in 180 µL of Buffer ATL (Qiagen). 300 µL of 0.1 mm zirconia/silica beads (BioSpec) were added followed by three cycles of vortex mixing for one minute and resting on ice for one minute. 20 µL of proteinase K (Qiagen) was then added and the samples were incubated for 3 hours at 56°C. Following incubation, 200 µL of Buffer AL was mixed into each sample and incubated for 10 minutes at 70°C. Finally, 200 µL of 100% ethanol was added to each sample. Following this, samples were applied to the QIAamp Mini spin columns and were spun (1 minute at 6000 x g) to bind to the column. Samples were washed with AW1 (Qiagen) followed by AW2 (Qiagen) using similar centrifugation settings. Lastly, 50 µL of nuclease-free water was added to each column and incubated for 1 minute at room temperature prior to elution via centrifugation.

#### *P. falciparum* Culture and RNA Extraction

*P. falciparum* 3D7 was cultured in human O+ red blood cells (RBCs) using standard RPMI 1640 medium supplemented with 0.5% Albumax II (Invitrogen), 2 mM L-glutamine, 50 mg L^−1^ hypoxanthine, 25 mM HEPES, 0.225% NaHCO3 and 10 mg mL^−1^ gentamicin. Cultures were incubated at 37°C and gassed with a sterile mixture of 5% O_2_, 5% CO_2_ and 90% N_2_. RNA was extracted from mixed *Plasmodium falciparum* 3D7 cultures at 1% parasitemia using two biological replicates. Cultures were washed with 1X PBS and transferred to a 50 mL tube. After centrifugation, the cell pellets were resuspended in TRIzol® LS Reagent (Invitrogen) and incubated at 37°C for 5 minutes. Chloroform was added, and the mixture was vortexed and centrifuged. The aqueous phase containing RNA was collected followed by RNA precipitation with isopropanol and incubation at −20°C overnight. The RNA was pelleted, washed with ethanol and air-dried prior to resuspension in RNase-free water. Genomic DNA contamination was removed by treating the RNA with 4 units of DNase I (NEB), followed by DNase inactivation with EDTA. RNA quality was assessed by agarose gel electrophoresis and serially diluted to 0.1 and 0.01% for further analysis.

#### HIV-1 RNA Extraction

Lysed HIV-1 viral samples were obtained from Prof. Otto Yang’s lab at UCLA. These samples were processed and the RNA was extracted using the Quick DNA/RNA™ MagBead Kit (Zymo Research) according to Supplementary Method M1.

### Dried Blood Spot Preparation and Elution

Dried Blood Spots (DBS) were prepared by spotting Qiagen FTA DBS Cards with whole blood samples containing spiked genetic material. Each spot was ~50 µL of whole blood, allowed to air dry for 4 hours. Spotted DBS cards were stored with desiccant at −20°C. For elution, each blood spot was cut out and placed in a tube with 500 µL of elution buffer (PBS + 0.05% Tween). Each tube was pulse vortexed 5 times followed by addition of 5 µL of Proteinase K (Zymo Research). Tubes were then incubated at 56°C for 1 hour and then heated at 95°C for 10 minutes. The eluates were then purified using the Quick DNA/RNA™ MagBead Kit (Zymo Research).

#### Extraction of Nucleic Acids from Sputum, Serum, Blood, and Dried Blood Spots

Nucleic acids from both contrived and patient samples were extracted using the Quick DNA/RNA™ MagBead Kit (Zymo Research) as described in Supplementary Method M1. Briefly, patient samples were mixed with DNA/RNA Shield™ in a 2:1 ratio with a final volume of 400 µL. This mix was supplemented with 8 µL of proteinase K (20 mg/mL) and incubated at 30°C for 30 minutes. Samples were then vortexed and centrifuged at 16,000 x g for 2 minutes to pellet debris. The supernatant was transferred to a new tube and mixed with an equivalent volume of 100% isopropanol. Following this pre-treatment, samples were purified and extracted as per Supplementary Method M1. The purified nucleic acids were eluted in nuclease-free water and immediately subjected to amplification.

### Testing of Contrived Samples

Purified nucleic acids and gBlocks corresponding to SNP detection targets were diluted and spiked into relevant sample matrices, including sputum, serum, whole blood, and dried blood spots, at final concentrations of 10^3^, 10^2^, 10^1^, 10^0^, and 10^−1^ copies µL^−1^. Each matrix and sample combination was prepared in 20 replicates and tested against the HTM panel of crRNAs. gBlocks corresponding to TB drug resistance targets and *M. tuberculosis* genomic DNA were evaluated in sputum, whole blood, and DBS. *P. falciparum* genomic DNA, HIV-1 genomic RNA, genomic material representing *P. ovale, P. vivax, P. malariae*, and HIV-2 were evaluated in whole blood, serum, and DBS. Purified nucleic acids were also used to generate contrived HIV-TB and HIV-*P. falciparum* coinfection samples.

Contrived coinfection samples were spiked to have combinations of either sample at final concentrations of 10^3^, 10^2^, 10^1^, and 10^0^ copies µL^−1^. All contrived samples underwent extraction using the Quick DNA/RNA™ MagBead Kit (Zymo Research), followed by CARMEN testing in technical duplicate using the Biomark HD. A previously designed pair of human RNase P primers and the corresponding crRNA were used as a positive control.

#### qPCR Testing of Contrived Samples

All samples were additionally evaluated by qPCR for comparison of detection sensitivity. RNA samples underwent a separate reverse-transcription step before qPCR as described above. qPCR was performed using 2X PowerUp SYBR Green Master Mix (Applied Biosystems) and the following thermocycling parameters: 2 minutes 50°C, 2 minutes at 95°C, and 40 cycles of 30 seconds of 95°C, 60 seconds of 58°C, and 60 seconds of 72°C. The same primer pools used for RT-PCR pre-amplification were used for RT-qPCR. RT-qPCR reactions were performed in triplicate with final reaction volumes of 10 µL, with tested samples comprising 10% of the total reaction volume. qPCR results were analyzed using Design and Analysis Software v2.7.0 (Applied Biosystems).

### Clinical patient samples

#### Ethics statement

All clinical specimens were obtained as de-identified specimens under the applicable institutional and commercial vendor protocols.

#### Healthy patient samples

Twenty unique, unfiltered serum samples from healthy human donors (10 male and 10 female) were obtained from BioIVT. Healthy whole blood samples were purchased from the UCLA Blood and Platelet Center. Serum and whole blood samples were aliquoted and spiked with genetic material from malaria and HIV sub-panel targets for contrived sample experiments. Healthy sputum was obtained as a kind donation from the Bertozzi Laboratory at Stanford University and was spiked with genomic material from the TB sub-panel for contrived sample experiments.

#### HIV-1 patient samples

Ten negative patient samples and 20 patient samples confirmed positive via RT-PCR for HIV-1 (with viral load ranging from 32 to 2,660,000 copies mL^−1^) were collected from the UCLA Clinical Microbiology Laboratory (IRB-25-2048). The plasma samples were heat inactivated before being stored at −80°C. Prior to CARMEN analysis, samples were lysed and nucleic acids were extracted using the Quick-DNA/RNA MagBead Kit (Zymo Research, Supplementary Method M1).

#### Malaria patient samples

Twenty patient samples confirmed positive via BinaxNOW Malaria and/or RT-PCR for malaria were purchased from Boca Biolistics. Of the twenty samples, 8 were serum and 12 were whole blood. The samples were shipped on dry ice and stored at −80°C. Prior to CARMEN analysis, the samples were processed by lysis and extraction using the Quick DNA/RNA™ MagBead Kit (Zymo Research, Supplementary Method M1).

#### Tuberculosis patient samples

30 extracted patient samples confirmed positive for *M. tuberculosis* via GeneXpert were provided by FIND Specimen Bank (Foundation for Innovative New Diagnostics, Geneva, Switzerland). The samples were shipped on dry ice and stored at −80°C. An additional nine patient plasma samples positive for both *M. tuberculosis* and HIV-1 were also purchased from FIND Specimen Bank. These 9 plasma samples were extracted using the Quick DNA/RNA™ MagBead Kit (Zymo Research) according to Supplementary Method M1.

### Data Analysis and Diagnostic Call Criteria

A Python script was used to parse, organize, and normalize raw fluorescence data from the Biomark HD. FAM reporter signal was normalized in each reaction chamber using the following formula:

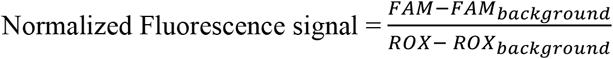

Diagnosis thresholds for CARMEN were set on a per crRNA basis. The threshold was set at the average NTC signal plus three times its standard deviation (SD) using the following formula:

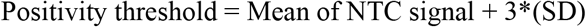

Samples were considered positive for a specific target if the normalized signal with its respective crRNA was above the threshold. Additionally, samples in qPCR were determined as positive/negative for a specific target based on the following cycle threshold (Ct) conditions:

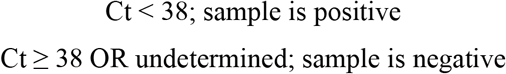

For SNP detection with the *inhA* and *katG* pairs of crRNAs, fluorescence ratios at 60 minutes between WT and MUT crRNAs for each gene are calculated and a diagnostic call for a dominant allele is made depending on whether a given ratio exceeds 2. For example, if *inhA* MUT/*inhA* WT for a sample containing TB (determined via *M. tuberculosis* IS6110 crRNA signal) exceeds 2, then it would indicate that the dominant *inhA* allele in that patient sample is the mutant allele as depicted in the following formula:

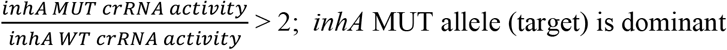

Clinical samples tested on CARMEN were determined to be positive or negative based on the positivity threshold calculation (as above). Positive Percent Agreement and Negative Percent Agreement were calculated using the following formula:

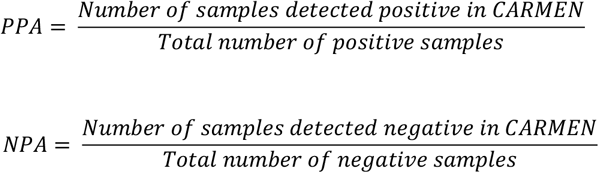

## Results

### Design and development of the HTM multiplex panel

We developed HTM CARMEN, a multiplex CRISPR-Cas13a panel designed to integrate pathogen identification, *Plasmodium* species differentiation, and Mtb isoniazid (INH) resistance-associated variant detection within a single assay (**Fig. 1A**). The panel comprises 11 diagnostic assays targeting *Plasmodium falciparum* (P*f*), *Plasmodium vivax* (P*v*), *Plasmodium ovale* (P*o*), *Plasmodium malariae* (P*m*), HIV-1, HIV-2, the *Mycobacterium tuberculosis* (Mtb) IS6110 insertion sequence, and wild-type and mutant alleles of the INH-resistance loci *inhA* promoter −777 C>T (also referred to as *fabG1* −15 C>T, Mtb *inhA*) and *katG* S315T/N (Mtb *katG*) (**Supplementary Table S1-S3**). For *Plasmodium* differentiation, crRNAs were intentionally designed against species-discriminating regions of the 18S rRNA locus while amplification primers were positioned within conserved flanking regions to enable simultaneous amplification of all four species (**Fig. 1B, Supplementary Table S4**). We additionally designed crRNAs against the common apicomplexan parasites *Babesia microti* (B*m*) and *Toxoplasma gondii* (T*g*) for use as specificity control assays (**Supplementary Table S2-S3, Supplementary Fig. S3A**). For each target, candidate crRNAs were designed based on publicly available genomic sequences and ranked using ADAPT, a machine learning-based design framework that predicts guide specificity and sensitivity^26^.

**Figure 1.**
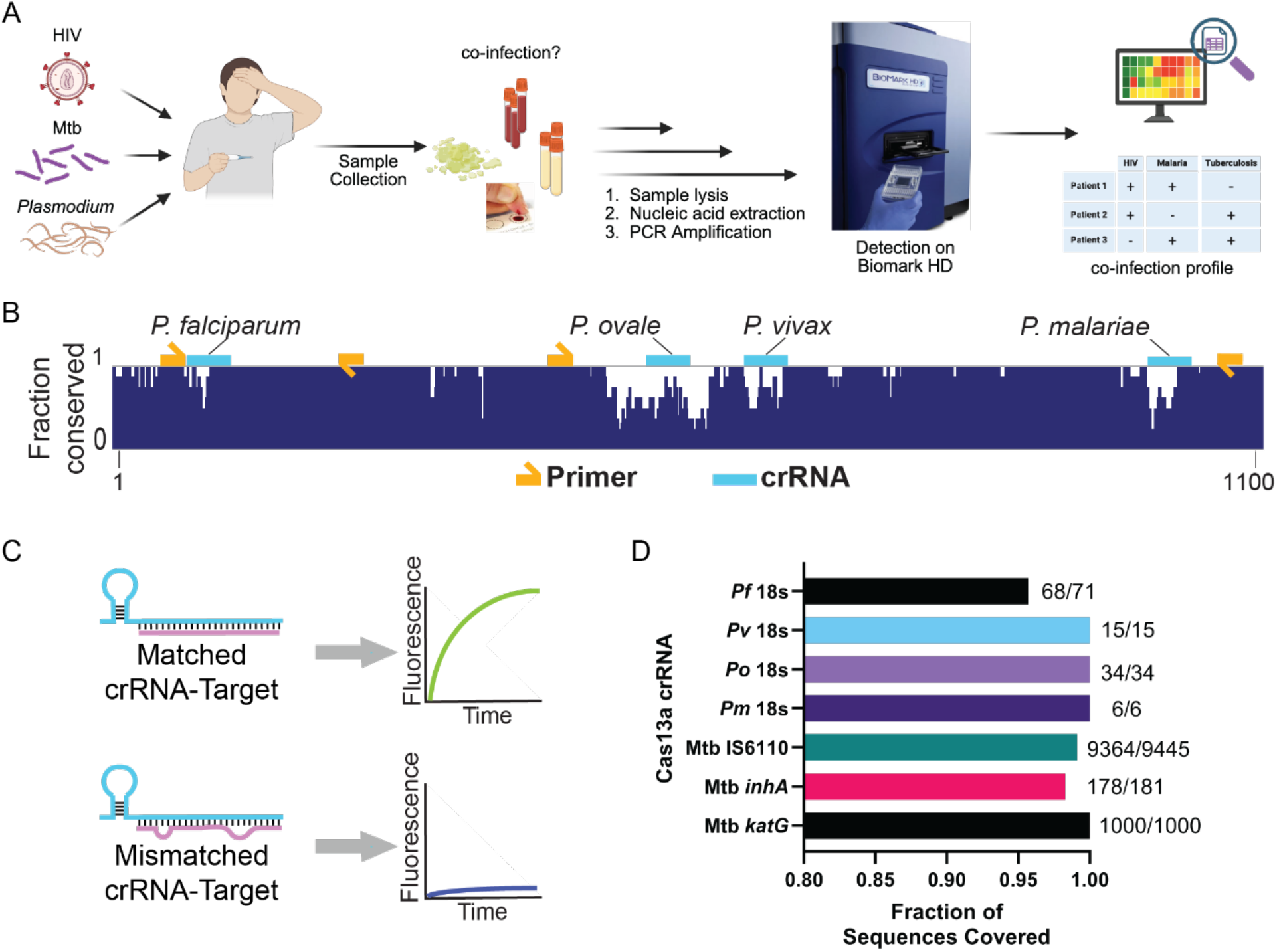
Design and workflow of the HTM CARMEN CRISPR diagnostic panel. **(A)** Schematic of the HTM CARMEN workflow, illustrating sample collection, nucleic acid extraction, target amplification, and CRISPR-Cas13a detection on the CARMEN platform, and multiplexed detection of HIV, *Plasmodium*, Mtb, and coinfections. Created in BioRender. Kamariza, M. (2026) https://BioRender.com/akepb0n **(B)** Design strategy for *Plasmodium* species differentiation. Sequence conservation across consensus 18S rRNA sequences from *P. falciparum, P. vivax, P. ovale*, and *P. malariae* are shown together with the positions of amplification primers and species-specific crRNAs. Primers were positioned within conserved regions, whereas crRNAs targeted species-discriminating regions identified using ADAPT. **(C)** Schematic of allele-specific crRNA detection for the Mtb *inhA* promoter −777 C>T and *katG* S315T/N loci. Cognate crRNA-target pairing promotes Cas13a activation and fluorescence generation, whereas a single-nucleotide mismatch reduces activation, enabling discrimination between wild-type and mutant alleles based on their relative fluorescence signals. **(D)** Predicted sequence coverage of the selected *Plasmodium* and Mtb assays across publicly available NCBI sequence datasets. Coverage represents the fraction of sequences predicted to support primer binding and crRNA activity. The number of sequences covered relative to the total evaluated is indicated for each assay.

For Mtb detection, we targeted the insertion sequence (IS) 6110, a multicopy element widely used as a sensitive molecular target for Mtb detection^28,29^. To simultaneously interrogate INH resistance, we designed allele-specific crRNAs targeting the *inhA* promoter −777 C>T and *katG* S315T/N loci. These two well-established genetic determinants associated with INH resistance in Mtb collectively account for a substantial fraction of INH-resistant isolates observed in the clinic and are routinely incorporated into molecular resistance detection assays^30–32^. Therefore, we designed *katG* and *inhA*-targeting crRNAs positioned across these specific loci, one crRNA targeting the wild-type (WT) and another targeting the mutant (MUT) gene sequence, to enable discrimination between WT and MUT *katG* or *inhA* Mtb alleles (**Fig. 1C**). These were designed using BADGERS-Cas13, a computational framework for iteratively mutating guides and predicting their activity against closely related allelic variants^27^. We adopted this dual-guide approach so that each genotype produces an independent fluorescent signal, rather than being inferred from attenuation of the wild-type signal. Finally, we included our previously validated HIV-1 and HIV-2 crRNAs to complete the HTM panel^25^.

We next assessed the predicted sequence coverage of the selected *Plasmodium* and Mtb crRNA assays against publicly available genomic sequence datasets (**Fig. 1D**). Across all assays, we found high predicted coverage ranging from 95% to 100% of publicly available genomic data. Specifically, we observed that the selected crRNAs are predicted to recognize 95.8% of P*f* 18S sequences (68/71 sequences), 100% of P*v* (15/15), 100% of P*o* (34/34), 100% of P*m* (6/6), 99.1% of Mtb IS6110 (9,364/9,445), 98.3% of *inhA* (178/181), and 100% of *katG* sequences (1,000/1,000). The available reference sets for P*v*, P*o*, and particularly P*m* were comparatively small, limiting the precision of these coverage estimates. Together, these analyses established a multiplex CRISPR panel predicted to simultaneously detect Mtb, HIV, and malaria, as well as profile *Plasmodium* species and INH resistance-associated Mtb variants.

### High specificity and SNP discrimination of HTM assays

We first evaluated the activity of the designed crRNAs against their cognate targets *in vitro* within individual SHERLOCK reactions^17^. Specifically, each amplified target, at decreasing concentrations, was incubated with their respective crRNA along with Cas13a in a detection reaction prior to fluorescence analysis (**Supplementary Fig. S1**). For all selected crRNAs, we observed rapid and robust fluorescence activation against their cognate targets, even at low concentrations, with minimal background signal. We then evaluated cross-reactivity in the multiplexed CARMEN format^25^. Genomic or synthetic material for each target included in the HTM panel was tested against every crRNA in the panel (**Fig. 2A, Supplementary Fig. S2**). We observed that across all assays, fluorescence activation remained largely restricted to intended cognate target-crRNA pairs, while Mtb genomic DNA produced the expected signal across IS6110 and the wild-type *inhA* and *katG* assays as these loci are present within the same Mtb genome. Because the WT and MUT targets at each resistance-associated locus differ by a single nucleotide, we quantified allele discrimination using paired *inhA* and *katG* crRNAs. We observed that at 10^4^ copies µL^−1^, each crRNA preferentially recognized its cognate allele, with mean on-target/off-target activity ratios of 13.69 ± 3.08 for *inhA* WT, 4.38 ± 0.92 for *inhA* MUT, 3.15 ± 0.39 for *katG* WT, and 3.99 ± 0.77 for *katG* MUT (**Fig. 2B, Supplementary Fig. S3B, Supplementary Table S5**). Thus, all four allele-specific crRNAs preferentially recognized their matched sequence, with greater discrimination observed for the *inhA* guides under these *in vitro* conditions.

**Figure 2.**
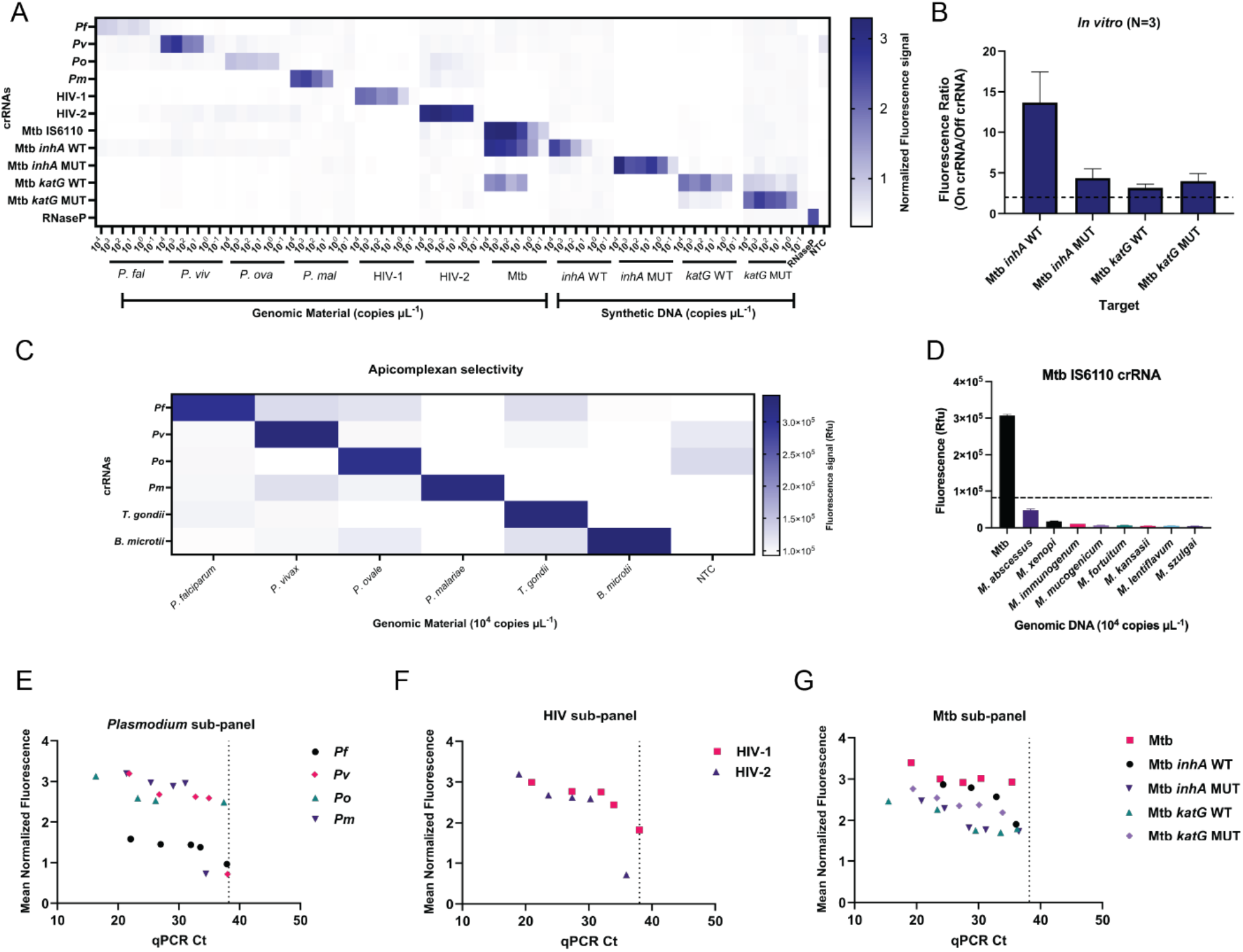
Analytical specificity and performance of the HTM CARMEN panel. (**A**) Multiplexed sensitivity and specificity of the HTM CARMEN assays under *in vitro* conditions. Heatmap shows normalized fluorescence at 60 min for the indicated genomic or synthetic targets across concentrations ranging from 10^4^ to 10^−1^ copies µL^−1^. (**B**) Allele discrimination of crRNAs targeting the *inhA* promoter −777 C>T and *katG* S315T/N loci. On-target/off-target fluorescence ratios were calculated at 60 min using wildtype Mtb genomic DNA or synthetic mutant gBlocks at 10^4^ copies µL^−1^. The dashed line indicates a ratio of 2, corresponding to no preferential allele recognition. Data are shown as mean ± SD from three independent biological replicates. (**C**) Reciprocal analytical specificity of the four *Plasmodium* species assays and the related apicomplexan controls *Toxoplasma gondii* and *Babesia microti*, tested at 10^4^ copies µL^−1^. (**D**) Specificity of Mtb IS6110 crRNA against Mtb and eight non-tuberculous mycobacterial species at 10^4^ copies µL^−1^. The dashed line indicates the assay positivity threshold defined as the no-target control mean plus three standard deviations. (**E-G**) Relationship between qPCR cycle threshold (Ct) and normalized CARMEN fluorescence across the *in vitro* target dilution series for (**E**) the *Plasmodium*, (**F**) HIV, and (**G**) Mtb sub-panels. The dotted vertical line denotes the qPCR Ct threshold of 38 cycles, above which qPCR results were considered undetermined. NTC = No-Target Control.

We further challenged HTM assay specificity using related organisms. In a reciprocal apicomplexan specificity matrix, the four *Plasmodium* assays preferentially detected their cognate species relative to the non-cognate *Plasmodium, Tg*, and *Bm* targets (**Fig. 2C**). We also evaluated the Mtb assays against genomic material from eight non-tuberculous mycobacteria (NTM) species. The Mtb IS6110, *inhA* WT, and *katG* WT crRNAs generated strong fluorescence signal against Mtb genomic DNA, whereas signal from the eight NTM species remained below the assay-specific positivity threshold (**Fig. 2D, Supplementary Fig. S3C-D**). Collectively, these experiments demonstrate the analytical selectivity of the HTM panel at pathogen, species, and single-nucleotide levels.

Finally, we compared normalized CARMEN fluorescence with qPCR cycle threshold (Ct) values across the same *in vitro* target dilution series. We found that normalized CARMEN fluorescence showed a consistent inverse relationship with qPCR Ct values across the *Plasmodium*, HIV, and Mtb sub-panels (**Fig. 2E-G**). When undetermined qPCR measurements (Ct values ≥38) were excluded, target-specific Pearson correlation coefficients ranged from −0.75 to −0.96, consistent with the expected inverse relationship between CARMEN fluorescence and qPCR Ct results across the tested concentration range (**Supplementary Table S6**).

### HTM CARMEN retains sensitive and selective detection in contrived samples

Having established analytical selectivity *in vitro*, we next asked whether HTM CARMEN retained its performance following sample processing in clinically relevant matrices. We evaluated sputum for Mtb targets, serum for HIV and malaria targets, and whole blood and dried blood spots (DBS) across multiple HTM sub-panels (**Fig. 3, Supplementary Fig. S4-S10**). In unspiked matrices, fluorescence remained low across the corresponding crRNAs, whereas contrived samples containing target material generated predominantly target-specific signal across the tested concentration series (**Supplementary Fig. S4**).

**Figure 3.**
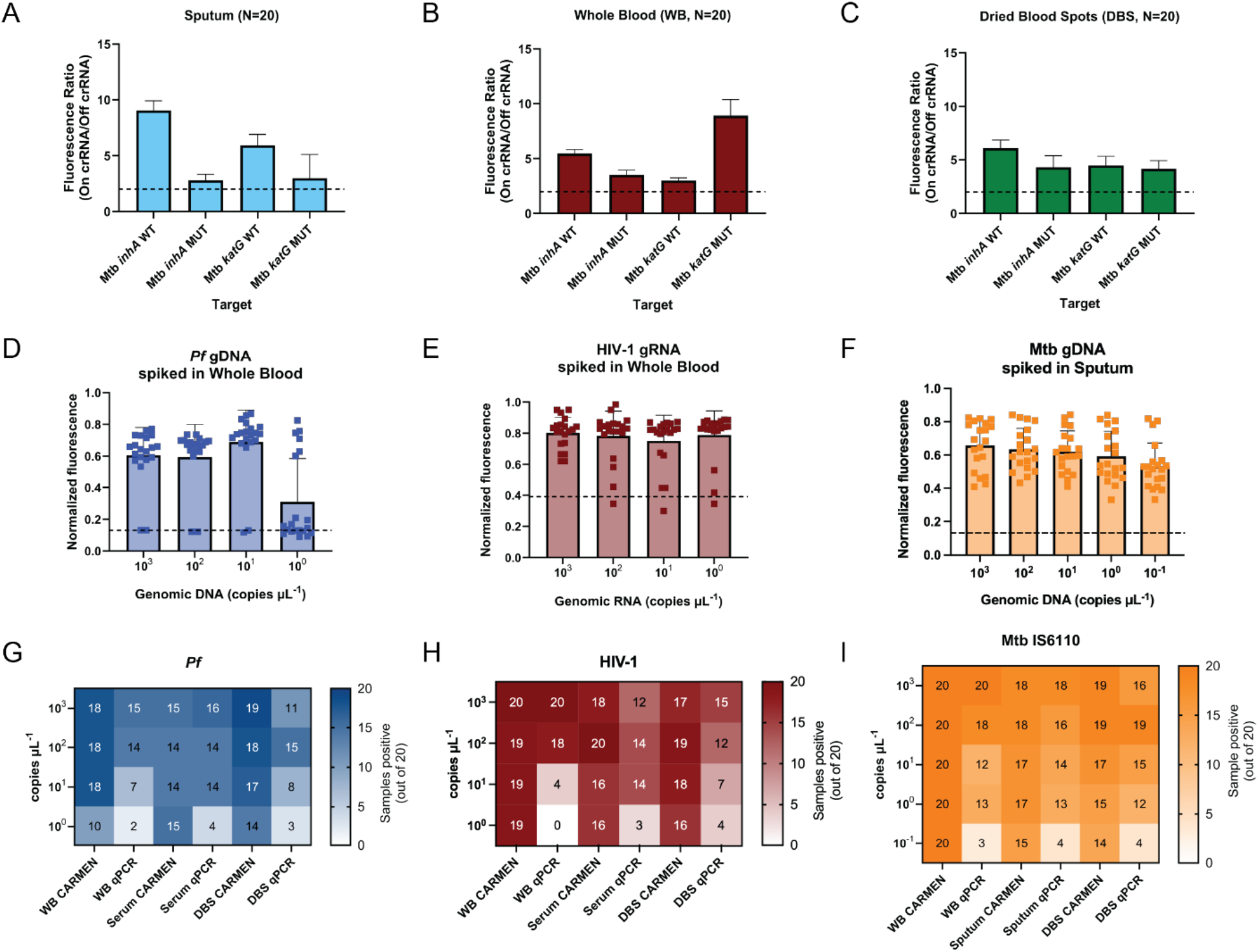
HTM CARMEN retains allele discrimination and sensitive detection across clinically relevant sample matrices. **(A-C)** Allele discrimination of Mtb *inhA* and *katG* crRNAs following testing in contrived **(A)** sputum, **(B)** whole blood, and **(C)** dried blood spot (DBS) samples. Activity ratios were calculated for each contrived sample at 10^4^ copies µL^−1^ as on-target/off-target normalized fluorescence at 60 min and are shown as mean ± SD from 20 samples. The dashed line indicates an activity ratio of 2, corresponding to no preferential allele recognition. **(D-F)** Detection of low-abundance targets across 20 contrived samples containing **(D)** *P. falciparum* genomic DNA in whole blood, **(E)** HIV-1 genomic RNA in whole blood, or **(F)** Mtb genomic DNA in sputum at the indicated input concentrations. Bars show mean normalized CARMEN fluorescence ± SD; individual points represent each contrived sample. Dashed lines indicate assay positivity thresholds. **(G-I)** Comparison of CARMEN and qPCR detection across target concentration and sample matrix for **(G)** *P. falciparum*, **(H)** HIV-1, and **(I)** Mtb IS6110. Heatmaps show the number of samples classified as positive out of 20 at each target concentration in whole blood, serum, and DBS for *P. falciparum* and HIV-1, and sputum, whole blood, and DBS for Mtb IS6110. Color scales reflect the number of samples classified as positive out of 20.

We first examined whether the single nucleotide discrimination established *in vitro* was retained following matrix processing. Across 20 contrived samples per matrix, cognate-allele preference was maintained for all four *inhA* and *katG* crRNAs in sputum, whole blood, and DBS (**Fig. 3A-C, Supplementary Fig. S5, Supplementary Table S7**). We observed mean on-target/off-target activity ratios ranged from 2.99 to 9.05 in sputum, 2.99 to 8.92 in whole blood, and 4.17 to 6.08 in DBS. Thus, allele discrimination was retained across all three matrices despite matrix variation.

We next quantified detection frequency across target concentrations using 20 contrived samples per condition. For P*f* genomic DNA in whole blood, CARMEN classified 18/20 samples as positive at 10^3^, 10^2^, and 10^1^ copies µL^−1^ and 10/20 at 10^0^ copies µL^−1^ (**Fig. 3D**). For HIV-1 genomic RNA in whole blood, CARMEN classified 20/20 samples as positive at 10^3^ copies µL^−1^ and 19/20 at each of 10^2^, 10^1^, and 10^0^ copies µL^−1^ (**Fig. 3E**). Mtb IS6110 detection in sputum remained positive in 20/20 samples across the full tested range from 10^3^ to 10^−1^ copies µL^−1^ (**Fig. 3F**). We further expanded our evaluation, which demonstrated detection across additional HTM targets and matrices, including serum, whole blood, DBS, and sputum (**Supplementary Figs. S6-S9**).

We then compared CARMEN and qPCR detection frequencies across the same target concentrations and matrices (**Fig. 3G-I**; **Supplementary Table S8**). Differences between the two methods were most apparent at low target inputs. At 10^1^ copies µL^−1^ in whole blood, CARMEN detected P*f* in 18/20 samples compared with 7/20 by qPCR, and HIV-1 in 19/20 compared with 4/20 by qPCR (**Fig. 3G,H**). For Mtb IS6110 in sputum, CARMEN detected 20/20 samples at 10^1^ copies µL^−1^ compared with 12/20 by qPCR, and 20/20 at 10^−1^ copies µL^−1^ compared with 3/20 by qPCR (**Fig. 3I**). As before, we found that matrix-specific CARMEN fluorescence measurements showed the expected inverse relationship with qPCR Ct for these representative pathogen-matrix combinations (**Supplementary Fig. S11**).

Across the broader target and matrix comparison, CARMEN generally classified an equal or greater number of low input samples as positive than qPCR, although the magnitude of the difference varied by target and matrix (**Fig. 3G-I, Supplementary Fig. S10)**. Together, these results demonstrate that HTM CARMEN retains target and allele discrimination following matrix processing and supports reproducible detection of low abundance targets across clinically relevant specimen types.

### Multiplexed detection of contrived HIV-malaria and HIV-TB coinfections

We next assessed whether HTM CARMEN can detect multiple co-occurring infections within the same multiplexed assay. We evaluated simultaneous detection of HIV-1 with either P*f* or Mtb in contrived coinfection samples across clinically relevant matrices. In whole blood containing P*f* and HIV-1, both pathogen targets generated fluorescence across their respective crRNAs, while the corresponding single-target samples produced the expected pathogen-specific signals (**Fig. 4A, Supplementary Fig. S12**). Similarly, in sputum containing Mtb and HIV-1, fluorescence activation was observed across the corresponding HIV-1 and Mtb assays, with minimal activation of non-cognate pathogen assays (**Fig. 4B, Supplementary Fig. S12**). These experiments established simultaneous target-selective detection in the presence of two pathogen inputs across a range of relative target concentrations.

**Figure 4.**
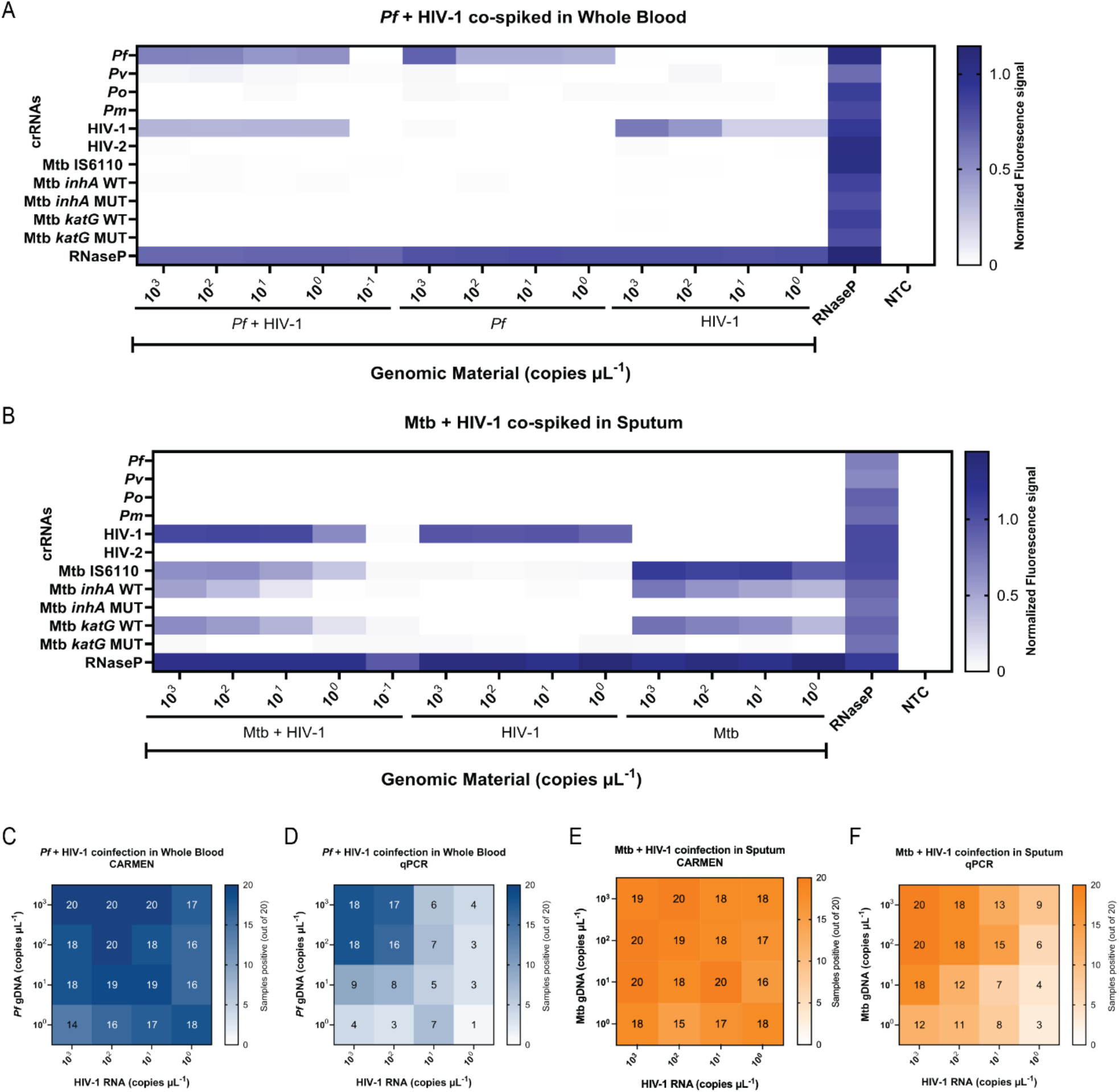
Multiplexed detection of contrived *Pf*-HIV-1 and Mtb-HIV-1 coinfections. **(A-B)** Multiplex CARMEN detection of **(A)** *Pf* and HIV-1 spiked simultaneously in whole blood and **(B)** Mtb and HIV-1 spiked simultaneously in sputum, at the indicated target concentrations. Heatmaps show normalized fluorescence at 60 min across the indicated HTM crRNAs for coinfected samples and corresponding singletarget controls across the indicated target concentrations. RNase P Positive Control and No-Target Control (NTC) are shown where indicated. **(C-F)** Detection frequencies across 20 contrived coinfection samples per concentration for **(C)** CARMEN and **(D)** qPCR detection of *Pf* and HIV-1 with their respective crRNAs in whole blood; and **(E)** CARMEN and **(F)** qPCR detection of Mtb and HIV-1 with their respective crRNAs in sputum. Heatmap values indicate the number of samples classified as positive out of 20 at each combination of HIV-1 and coinfecting pathogen concentration. Color scales reflect the number of samples classified as positive out of 20.

We next evaluated detection frequency across 20 contrived coinfection samples at each combination of pathogen concentrations and compared CARMEN with qPCR (**Fig. 4C-F, Supplementary Fig. S12**). For P*f* and HIV-1 coinfections, CARMEN maintained high detection frequencies across much of the concentration matrix, including conditions in which one or both targets were present at low input (**Fig. 4C**). Differences between CARMEN and qPCR became more pronounced toward the lower end of the tested concentration range (**Fig. 4C-D**). When both targets were present at 10^1^ copies µL^−1^, CARMEN detected P*f* in 19/20 samples and HIV-1 in 18/20, compared with 8/20 and 9/20 by qPCR, respectively. At 10^0^ copies µL^−1^ for both targets, CARMEN detected P*f* in 18/20 samples and HIV-1 in 14/20, compared with 3/20 and 4/20 by qPCR, respectively. Similarly, CARMEN simultaneously detected Mtb and HIV-1 across the contrived sputum coinfection matrix (**Fig. 4E, Supplementary Fig. S12**). When both targets were present at 10^1^ copies µL^−1^, CARMEN detected Mtb in 20/20 samples and HIV-1 in 20/20, compared with 12/20 and 18/20 by qPCR, respectively (**Fig. 4E-F**). At 10^0^ copies µL^−1^ for both targets, CARMEN detected Mtb and HIV-1 in 18/20 samples each, compared with 11/20 and 12/20 by qPCR, respectively. Across the broader concentration matrix, detection frequency varied with the relative abundance of the two pathogen targets.

Together, these experiments demonstrate simultaneous detection of two pathogen targets within the multiplexed HTM CARMEN workflow across a range of target concentrations. The increased detection frequency observed for low-input HIV-1 and P*f* coinfections further indicates that CARMEN multiplexing did not preclude detection in conditions where one or both pathogen targets approached the lower end of the tested concentration range. Importantly, in the low-input conditions tested, CARMEN frequently detected a greater proportion of contrived coinfection samples than qPCR.

### Clinical detection of HIV, malaria, tuberculosis, and HIV-TB coinfection

Having established analytical performance in contrived matrices, we next evaluated HTM assays using nucleic acids extracted from cultured pathogen material. CARMEN detected P*f* across the tested parasitemia range of 1% to 0.01%, HIV-1 RNA across input concentrations from 10^7^ to 10^0^ copies µL^−1^, and Mtb across the tested bacterial concentrations of 10,000, 100 and 1 CFU mL^−1^ (**Supplementary Fig. S13A-C**). Across these experiments, CARMEN fluorescence showed the expected inverse relationship with qPCR Ct (**Supplementary Fig. S13D-F**).

We next evaluated HTM CARMEN using clinical specimens with confirmed positivity for HIV, malaria, or Mtb infection. The complete multiplex panel was applied to each specimen, enabling simultaneous interrogation of the corresponding pathogen targets (**Fig. 5**). Among 20 confirmed-positive malaria specimens, CARMEN detected malaria in 19/20 (95%), while all 10 confirmed-negative specimens were classified as negative (**Fig. 5A-B, Supplementary Fig. S14**). Similarly, CARMEN detected HIV-1 in 19/20 (95%) confirmed-positive specimens, with 10/10 confirmed-negative specimens classified as negative (**Fig. 5C-D, Supplementary Fig. S14**).

**Figure 5.**
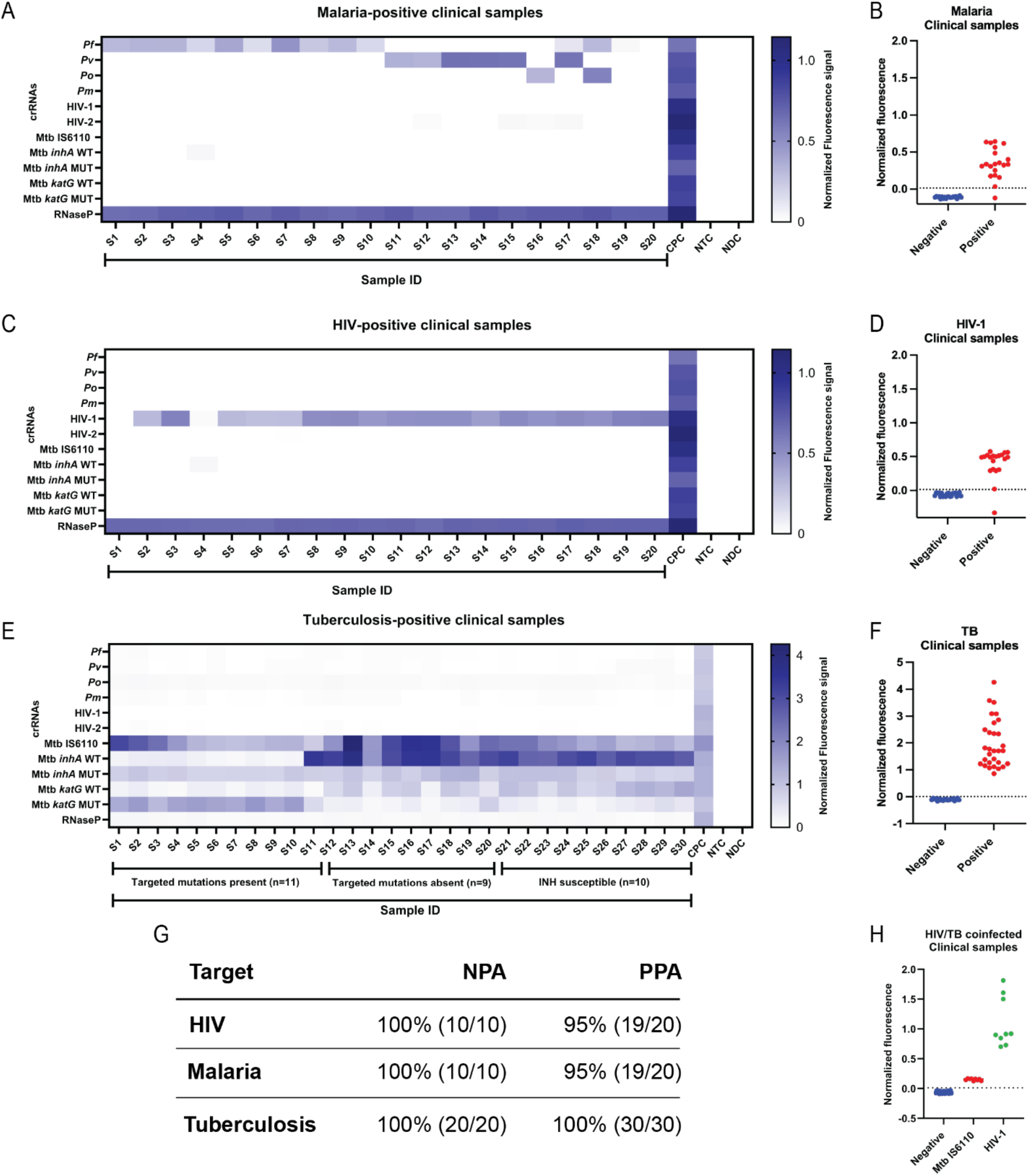
Detection of malaria, HIV-1, TB, and HIV-TB coinfection in clinical specimens. **(A-B)** Analysis of malaria clinical specimens (S1-20), showing **(A)** normalized CARMEN fluorescence across the indicated HTM crRNAs for 20 confirmed-positive samples and **(B)** normalized fluorescence in confirmed-positive and confirmed-negative specimens. **(C-D)** Analysis of HIV-1 clinical specimens, showing **(C)** normalized CARMEN fluorescence across the indicated HTM crRNAs for 20 confirmed-positive specimens and **(D)** normalized fluorescence in confirmed-positive and confirmed-negative specimens. **(E-F)** Analysis of TB clinical specimens, showing **(E)** normalized CARMEN fluorescence across the Mtb sub-panel for 30 GeneXpert-positive specimens and **(F)** Mtb IS6110 assay fluorescence in confirmed-positive and confirmed-negative specimens. **(G)** Positive percent agreement (PPA) and negative percent agreement (NPA) between CARMEN classification and reference infection status for malaria, HIV, and TB. **(H)** Normalized fluorescence for Mtb IS6110 and HIV-1 in nine confirmed-positive HIV-TB coinfected clinical specimens compared with confirmed-negative specimens. For all plotted graphs, individual points represent clinical specimens, and the dashed lines indicate the corresponding positivity threshold. Negative normalized fluorescence values reflect signal below the background reference used for normalization. CPC = combined positive control. NTC = No-Target Control. NDC = No Detection Control.

We next evaluated 30 clinical specimens confirmed positive for Mtb. CARMEN detected Mtb IS6110 in 30/30 (100%) confirmed-positive specimens, while 20/20 confirmed-negative specimens remained negative for IS6110 (**Fig. 5E-F, Supplementary Fig. S14**). Because the complete Mtb sub-panel was run simultaneously, the clinical specimens were also interrogated with the allele-specific *inhA* and *katG* assays. Reference mutation annotations supplied with the FIND specimens identified 11 of 30 specimens as carrying both CARMEN-targeted resistance-associated variants, *inhA* promoter −777 C>T and *katG* S315T/N. CARMEN classified all 11 specimens as carrying the corresponding mutant alleles, concordant with the supplied reference mutation annotations (**Fig. 5E**). The remaining specimens were either INH-susceptible (10/30) or included isolates with resistance-associated variants (9/30) outside the two alleles interrogated by the present panel, underscoring that the *inhA* −777 C>T and *katG* S315T/N assays capture a defined subset of the genetic diversity associated with INH resistance.

Across the three single-pathogen clinical cohorts, HTM CARMEN showed high agreement with reference infection status (**Fig. 5G**). Positive percent agreement (PPA) was 95% (19/20) for malaria, 95% (19/20) for HIV-1, and 100% (30/30) for Mtb IS6110, while negative percent agreement (NPA) was 100% for malaria (10/10), HIV-1 (10/10) and Mtb IS6110 (20/20). These results demonstrate consistent classification of confirmed-positive and confirmed-negative clinical specimens across the three pathogen classes tested.

Finally, we tested nine clinical specimens confirmed positive for both HIV-1 and Mtb to determine whether the multiplexed assay could identify both pathogens simultaneously. CARMEN detected HIV-1 and Mtb IS6110 concurrently in all nine specimens (**Fig. 5H, Supplementary Fig. S14**). Thus, the simultaneous pathogen detection observed in contrived coinfections extended to clinical HIV-TB coinfection specimens.

Together, these clinical experiments demonstrate high agreement between HTM CARMEN and reference infection status across the HIV-1, malaria, and Mtb specimens tested, concordance with supplied reference annotations for the targeted Mtb resistance-associated variants, and simultaneous detection of HIV-1 and Mtb in clinical coinfection samples. Combined with the analytical and contrived-matrix experiments, these results highlight the potential of HTM CARMEN for integrated pathogen, resistance-associated variant and coinfection detection within a single multiplexed workflow.

## Discussion and Conclusion

HIV, tuberculosis (TB), and malaria are managed through largely disease-specific diagnostic pathways despite substantial geographic overlap and clinically important coinfection^1–3,7,8,12,13^. Here, we developed HTM CARMEN as a multiplex CRISPR-Cas13a diagnostic framework that integrates detection of HIV-1 and HIV-2, four *Plasmodium* species, and *Mycobacterium tuberculosis* (Mtb) alongside resistance-associated variants in *inhA* and *katG*. Across analytical experiments *in vitro* and with clinically relevant sample matrices, the HTM panel retained target selectivity and single-nucleotide discrimination while enabling simultaneous detection of HIV, *Plasmodium*, and Mtb. Using confirmed-positive clinical specimens, our HTM panel showed 95% positive percent agreement (PPA) for malaria (19/20), 95% PPA for HIV-1 (19/20) and 100% PPA for Mtb IS6110 (30/30), with 100% negative percent agreement (NPA) for each target in the confirmed-negative specimens tested. Both HIV-1 and Mtb were also detected in all nine confirmed-positive HIV-TB coinfection specimens. Together, these findings establish the feasibility of integrating pathogen detection, species differentiation, resistance-associated variant interrogation and coinfection detection within a single programmable molecular testing platform.

The potential value of the HTM CARMEN approach lies not simply in multiplexing across viral, bacterial and parasitic pathogens, but in organizing that capability around diagnostic needs in co-endemic regions. Indeed, in settings where these infections co-circulate, identification of one pathogen does not exclude another, yet diagnostic pathways commonly evaluate each disease independently. This mismatch is particularly consequential for HIV-TB, where coinfection alters disease presentation and can complicate conventional sputum-based diagnosis. It also applies more broadly where malaria, HIV, and TB coexist within the same populations^7,8,14^. HTM CARMEN instead interrogates these infections in parallel. The 192 by 24 microfluidic architecture combinatorially tests up to 24 assays against 192 samples, corresponding to 4,608 assay-sample reactions per integrated fluidic circuit^23,25^. In its current form, this architecture is therefore more naturally positioned as a high-throughput strategy for centralized or regional molecular laboratories than as a replacement for decentralized rapid tests. Its greatest advantage is the ability to consolidate information that would otherwise require multiple disease-specific molecular workflows into a common analytical framework.

The contrived coinfection experiments provide an important test of this system. Simultaneous detection was maintained across varying concentrations of *P. falciparum*/HIV-1 and Mtb/HIV-1, including conditions in which the relative abundance of the two targets differed. At lower target inputs, HTM CARMEN frequently classified more contrived samples as positive than qPCR under the conditions tested, with particularly pronounced differences in the *P. falciparum*/HIV-1 concentration matrix. These experiments show that multiplex interrogation remained effective when two pathogen targets were simultaneously present and across a range of relative target abundances. The subsequent detection of both HIV-1 and Mtb in all nine confirmed-positive clinical HIV-TB coinfection specimens provides an initial bridge between this analytical capability and clinical coinfection detection. Larger studies spanning a broader range of HIV viral loads, TB bacillary burdens, disease phenotypes and specimen types will be required to establish whether integrated testing improves coinfection ascertainment relative to current diagnostic algorithms.

An additional feature of HTM CARMEN is that multiplexing extends beyond pathogen presence or absence. Species-level malaria identification was incorporated by combining amplification of conserved regions with species-discriminating *Plasmodium* crRNAs. This capability may be relevant where species composition informs treatment, relapse risk, or surveillance^33,34^. Similarly, paired crRNAs targeting the *inhA* promoter −777 C>T and *katG* S315T/N loci enabled single nucleotide discrimination between wild-type and mutant Mtb targets. Allelic preference was retained following processing in sputum, whole blood and dried blood spots, indicating that the discrimination observed *in vitro* was maintained across the tested matrices.

Integrating resistance-associated variant interrogation with pathogen detection is particularly relevant for TB because isoniazid (INH)-resistance can occur in rifampicin-susceptible disease and may therefore be missed by diagnostic strategies centered primarily on rifampicin resistance^15,32^. The present panel addresses two common INH-resistance-associated variants and should consequently be viewed as proof of principle for integrated resistance genotyping. Future iterations could extend resistance coverage to *rpoB* variants associated with rifampicin resistance, *gyrA/gyrB* variants associated with fluoroquinolone resistance, and additional determinants of resistance to first- and second-line TB drugs^32^.

Several limitations constrain clinical conclusions that can be drawn from the present study. First, the clinical cohorts were relatively small and consisted of reference-characterized specimens rather than patients prospectively enrolled from an intended-use population. The PPA and NPA observed here therefore quantify agreement within the specimens tested. Prospective evaluation in geographically diverse cohorts will be required to capture variation in pathogen burden, strain diversity, disease stage, treatment status and specimen quality. Such studies should also evaluate whether simultaneous testing materially changes diagnostic yield, time to appropriate treatment or recognition of coinfection compared with current sequential diagnostic pathways.

The resistance component requires a distinct next stage of validation. Reference mutation annotations supplied with the clinical Mtb specimens identified *inhA* promoter −777 C>T and *katG* S315T/N in 11 of 30 specimens, and CARMEN allele calls were concordant with these annotations. However, we did not have access to the underlying genotyping data or paired phenotypic drug-susceptibility results. Further clinical validation will therefore require comparison with independently determined genotypes and phenotypic drug-susceptibility testing. Moreover, *inhA* −777 C>T and *katG* S315T/N capture only a subset of the genetic diversity underlying INH resistance^30,32^. This limitation was evident in the Mtb clinical cohort: Out of the remaining 19 Mtb specimens, 10 were susceptible to INH, whereas the other nine were INH resistant and harbored resistance-associated variants outside the loci interrogated by the current panel. Thus, although HTM CARMEN accurately distinguished the targeted alleles, the present resistance sub-panel cannot capture the broader spectrum of INH-resistance-associated variation. A clinically actionable resistance panel would require broader variant coverage and prospective validation of genotype-to-phenotype classification. Reliance on Mtb IS6110 also introduces a recognized limitation of the current Mtb detection strategy. Although its multicopy nature supports sensitive detection, IS6110 copy number varies among Mtb strains and isolates with few or absent copies can occur, creating the potential for false-negative results in assays relying exclusively on this target^35,36^. Notably, the prevalence of IS6110-negative strains is geographically variable, reinforcing the value of target redundancy rather than assuming a universal frequency. Incorporating an additional conserved Mtb target in future panel iterations could provide redundancy against this source of false-negative detection.

The malaria component is also intentionally focused. The current implementation targets *P. falciparum, P. vivax, P. ovale* and *P. malariae*, although future panel expansion could incorporate zoonotic species such as *P. knowlesi*^34,37,38^. More broadly, computational sequence coverage and analytical testing against available genomic material cannot substitute for empirical validation against geographically diverse parasite, viral and bacterial populations. Indeed, sequence diversity can alter both primer amplification and crRNA recognition. Previous CARMEN resistance testing, for example, showed reduced agreement when HIV assays designed against subtype B sequences were applied to subtype G specimens, with improved concordance after accounting for primer and guide mismatches^22^. Nevertheless, the programmable nature of CRISPR detection provides an excellent mechanism for updating panel content as pathogen diversity and surveillance priorities change.

Implementation will also depend on more than analytical performance. The current workflow requires nucleic acid extraction, reverse transcription where appropriate, multiplex amplification and operation of the Biomark microfluidic platform. These requirements make the present implementation better suited to centralized or regional laboratories than to decentralized point-of-care testing. Previous development of microfluidic CARMEN demonstrated that automation and workflow consolidation could reduce processing time and manual handling, including reducing the workflow from more than 8 h to less than 5 h^23^. Whether consolidation of multiple disease-specific tests offsets the requirements for instrumentation, technical expertise, reagent supply and data analysis will require formal workflow and health-economic evaluation. Time-to-result and hands-on time were not formally benchmarked in the present study and should be evaluated alongside diagnostic performance in future implementation studies.

A further translational challenge is specimen selection. Although HTM CARMEN performed well across serum, whole blood, sputum and dried blood spots in contrived experiments, this does not establish that a single specimen type is optimal for simultaneous clinical detection of HIV, TB and malaria. Indeed, current diagnostic pathways use different specimen types and testing modalities according to pathogen, disease presentation and intended clinical question^5,33,39,40^. Therefore, prospective studies in high-burden settings represent an important next step. These future studies should evaluate locally collected specimens across relevant pathogen burdens and disease phenotypes, establish clinical performance against standardized reference testing, and determine whether integrated testing improves recognition of coinfection or alters clinical management.

Ultimately, HTM CARMEN is complementary to existing diagnostic algorithms. WHO guidance supports rapid molecular diagnostics for initial TB testing in appropriate populations, including Xpert MTB/RIF Ultra and Truenat-class assays, with molecular drug-resistance testing incorporated into subsequent diagnostic pathways^39^. For malaria, microscopy and rapid diagnostic tests remain the primary tools for clinical diagnosis, whereas nucleic acid amplification enables detection of lower burden infections but is not currently recommended by WHO for routine clinical management^33,34^. HIV diagnosis likewise relies primarily on established serological testing strategies, with molecular testing occupying defined diagnostic and monitoring roles^40^. Within this landscape, HTM CARMEN offers a centralized molecular assay that can interrogate HIV, TB and malaria targets concurrently while providing species- and resistance-associated sequence information.

Overall, HTM CARMEN provides a proof of concept for organizing molecular infectious-disease testing around co-endemic infections and coinfections. The present study establishes analytical integration across viral, bacterial and parasitic targets, demonstrates species- and allele-level discrimination across multiple sample matrices, and provides initial clinical evidence for malaria, HIV and TB detection together with simultaneous HIV/Mtb detection in coinfected specimens. The next challenge is to determine whether this breadth translates into improved diagnostic yield, treatment selection or surveillance efficiency in populations for which integrated testing is intended. If validated prospectively, HTM CARMEN could complement disease-specific rapid diagnostics by providing regional laboratories with a common molecular framework for simultaneously detecting multiple infections and selected resistance genotypes.

## Data Availability

All data produced in the present study are available upon reasonable request to the authors.

## Acknowledgements

We thank Bradley lab from UCLA for *T. gondii* genomic DNA samples. We thank Bertozzi lab for sputum samples from Stanford University. We gratefully acknowledge the FIND Specimen Bank / FIND Integrated Biobanks and participating clinical and laboratory staff for providing the biological samples used in this study. We extend our sincere thanks to the individual donors who made this research possible. We acknowledge the Boca Biolistics for providing characterized clinical samples. We are grateful for the support from the *UCLA-CDU CFAR grant AI152501 and the Pendleton Trust* to O.Y. This research was supported by startup funds and a UCLA Hellman Fellowship to M.K.

## Author contributions

M.K. conceived, designed, and supervised the study. A.S.J. and A.S. performed most of the experiments with J.Y., L. A. S. and C. S. providing technical support. A. N. and K. G. L. R. provided the extracted *P. falciparum* genomic material. A. B. and O. Y. provided lysed HIV-1 viral particles. O. G. provided HIV clinical samples. A.S.J., A.S., and M.K. analyzed all the data, wrote and edited the manuscript, which was approved by all authors.

## Declaration of generative AI and AI-assisted technologies

During the preparation of this work, the authors used ChatGPT and Claude to assist with data review and to refine text. The authors reviewed and edited all AI-assisted outputs and take full responsibility for the results and content of the manuscript.

