## Supplementary Information for "Integrated CRISPR-based detection of HIV, tuberculosis, malaria, and resistance-associated variants"

### I. Supporting Schemes and Tables

| Pathogen name | Acronym | Gene target |
|---|---|---|
| *Plasmodium falciparum* | $P_f$ | *18s* |
| *Plasmodium vivax* | $P_v$ | *18s* |
| *Plasmodium ovale* | $P_o$ | *18s* |
| *Plasmodium malariae* | $P_m$ | *18s* |
| Human Immunodeficiency Virus 1 | HIV-1 | *Pol protein* |
| Human Immunodeficiency Virus 2 | HIV-2 | *Gag polyprotein* |
| *Mycobacterium tuberculosis* IS6110 | Mtb IS6110 | IS 6110 |
| *Mycobacterium tuberculosis inhA* | Mtb *inhA* WT | *inhA* |
| *Mycobacterium tuberculosis inhA* -777C>T | Mtb *inhA* MUT | *inhA* |
| *Mycobacterium tuberculosis katG* S315 | Mtb *katG* WT | *katG* |
| *Mycobacterium tuberculosis katG* S315T/N | Mtb *katG* MUT | *katG* |

**Supplementary Table S1.** Genes targeted by HTM panel crRNAs and their nomenclature.

| **Targets** | **crRNA sequence** |
|---|---|
| $\boldsymbol{P_f}$ | GAUUUAGACUACCCCAAAAACGAAGGGGACUAAAACGAUACACACUAAAUAAAAUAAAUUUAUU |
| $\boldsymbol{P_v}$ | GAUUUAGACUACCCCAAAAACGAAGGGGACUAAAACCAAAGAAAGUCCUUAAAAAGAAUCAUUU |
| $\boldsymbol{P_o}$ | GAUUUAGACUACCCCAAAAACGAAGGGGACUAAAACUUUGGAUAAGGAAUGCAAAGAGCAGAUA |
| $\boldsymbol{P_m}$ | GAUUUAGACUACCCCAAAAACGAAGGGGACUAAAACGAAGGAAGCUAUCUAAAAGAAACACUCA |
| **HIV-1** | GAUUUAGACUACCCCAAAAACGAAGGGGACUAAAACAUACUGUAUCAUCUGCUCCUGUAUCUAA |
| **HIV-2** | GAUUUAGACUACCCCAAAAACGAAGGGGACUAAAACUUUAGGGUUCGGGGACUCAGCGGCACAU |
| **Mtb IS6110** | GAUUUAGACUACCCCAAAAACGAAGGGGACUAAAACGAGGGCAUCGAGGUGGCCAGAUGCACCG |
| **Mtb *inhA* WT** | GAUUUAGACUACCCCAAAAACGAAGGGGACUAAAACCACCCCGACAAGCUAUCGUCUCGCCGCG |
| **Mtb *inhA* MUT** | GAUUUAGACUACCCCAAAAACGAAGGGGACUAAAACCCUAUCAGCUCGCCGCGUCCGGGCCGAA |
| **Mtb *katG* WT** | GAUUUAGACUACCCCAAAAACGAAGGGGACUAAAACUACAACCUCGAUGCCCCUCGUGAUCGCG |
| **Mtb *katG* MUT** | GAUUUAGACUACCCCAAAAACGAAGGGGACUAAAACUGGUUAUCGCGUCCUUACCCGUUCCGGU |
| ***T .gondii*** | GAUUUAGACUACCCCAAAAACGAAGGGGACUAAAACGCAGACCGAAGUCAACGCGAUCCGUUCG |
| ***B. microti*** | GAUUUAGACUACCCCAAAAACGAAGGGGACUAAAACACUGACGACCUCCAAUCUCUAGUCGGCA |

**Supplementary Table S2.** crRNA sequences designed and selected for the HTM panel detection.

| **Pathogen Name** | **crRNA (ssDNA template sequence 5' – 3')** |
|---|---|
| *Plasmodium falciparum* | AATAAATTTATTTTATTTAGTGTGTATCGTTTTAGTCCCCTTCGTTT<br>TTGGGGTAGTCTAAATCCCCTATAGTGAGTCGTATTAATTTC |
| *Plasmodium vivax* | AAATGATTCTTTTTAAGGACTTTCTTTGGTTTTAGTCCCCTTCGTTT<br>TTGGGGTAGTCTAAATCCCCTATAGTGAGTCGTATTAATTTC |
| *Plasmodium ovale* | TATCTGCTCTTTGCATTCCTTATCCAAAGTTTTAGTCCCCTTCGTTT<br>TTGGGGTAGTCTAAATCCCCTATAGTGAGTCGTATTAATTTC |
| *Plasmodium malariae* | TGAGTGTTTCTTTTAGATAGCTTCCTTCGTTTTAGTCCCCTTCGTTTT<br>TGGGGTAGTCTAAATCCCCTATAGTGAGTCGTATTAATTTC |
| Human Immunodeficiency Virus 1 | TTAGATACAGGAGCAGATGATACAGTATGTTTTAGTCCCCTTCGTT<br>TTTGGGGTAGTCTAAATCCCCTATAGTGAGTCGTATTAATTTC |
| Human Immunodeficiency Virus 2 | ATGTGCCGCTGAGTCCCCGAACCCTAAAGTTTTAGTCCCCTTCGTT<br>TTTGGGGTAGTCTAAATCCCCTATAGTGAGTCGTATTAATTTC |
| *Mycobacterium tuberculosis* | CGGTGCATCTGGCCACCTCGATGCCCTCGTTTTAGTCCCCTTCGTTT<br>TTGGGGTAGTCTAAATCCCCTATAGTGAGTCGTATTAATTTC |
| *Mycobacterium tuberculosis inhA* -777C | CGCGGCGAGACGATAGCTTGTCGGGGTGGTTTTAGTCCCCTTCGTT<br>TTTGGGGTAGTCTAAATCCCCTATAGTGAGTCGTATTAATTTC |
| *Mycobacterium tuberculosis inhA* -777C>T | TTCGGCCCGGACGCGGCGAGCTGATAGGGTTTTAGTCCCCTTCGTT<br>TTTGGGGTAGTCTAAATCCCCTATAGTGAGTCGTATTAATTTC |
| *Mycobacterium tuberculosis katG* S315 | CGCGATCACGAGGGGCATCGAGGTTGTAGTTTTAGTCCCCTTCGTT<br>TTTGGGGTAGTCTAAATCCCCTATAGTGAGTCGTATTAATTTC |
| *Mycobacterium tuberculosis katG* S315T/N | ACCGGAACGGGTAAGGACGCGATAACCAGTTTTAGTCCCCTTCGTT<br>TTTGGGGTAGTCTAAATCCCCTATAGTGAGTCGTATTAATTTC |

**Supplementary Table S3.** crRNA templates ordered as ssDNA for *in vitro* transcription via T7 RNA Polymerase.

| Targets | Forward Primer | Reverse Primer |
|---|---|---|
| $P_f$ | TAATACGACTCACTATAgggTTAAGGAATTATAACAAAGAAGTAACACG | CCTGCTGCCTTCCTTAGATG |
| $P_v$ | TAATACGACTCACTATAgggTTGTTGCAGTTAAAACGCTCG | ATACTCGCCCCAGAACCC |
| $P_o$ | TAATACGACTCACTATAgggTTGTTGCAGTTAAAACGCTCG | ATACTCGCCCCAGAACCC |
| $P_m$ | TAATACGACTCACTATAgggTTGTTGCAGTTAAAACGCTCG | ATACTCGCCCCAGAACCC |
| **HIV-1** | gaaatTAATACGACTCACTATAgggCTTTGGCAACGAC CCCTC | CCCCTATCATTTTTGGTTTCCATT |
| **HIV-2** | gaaatTAATACGACTCACTATAgggTGCACGCAGAAGAGAAAGTG | GCCCCGAACTTCTTTTCCTC |
| **Mtb IS6110** | gaaatTAATACGACTCACTATAggggtaggcgtcggtgacaaag | CGAACTCAAGGAGCACATCA |
| **Mtb *inhA* WT** | gaaatTAATACGACTCACTATAgggAGTCACACCGACAAACGTC | GGATACGAATGGGGGTTTGG |
| **Mtb *inhA* MUT** | gaaatTAATACGACTCACTATAgggAGTCACACCGACAAACGTC | GGATACGAATGGGGGTTTGG |
| **Mtb *katG* WT** | TAATACGACTCACTATAgggGTCACACTTTCGGTAAGACCC | GTCCTTGGCGGTGTATTGC |
| **Mtb *katG* MUT** | TAATACGACTCACTATAgggGTCACACTTTCGGTAAGACCC | GTCCTTGGCGGTGTATTGC |
| ***T .gondii*** | TAATACGACTCACTATAGAGATACAGAACCAACCCACCT | CCTGCTGCCTTCCTTAGATG |
| ***B. microti*** | TAATACGACTCACTATAGACCGTCGTAATCCTAACCATAAAC | ATTTCTCTCAAGGTGCTGAAGG |
| **T7 Promoter** | GAAATTAATACGACTCACTATAGGG | N/A |

**Supplementary Table S4:** Primer sequences designed to amplify the gene of interest for all the HTM panel targets.

| Target | On-target crRNA mean normalized fluorescence ± SD | Off-target crRNA mean normalized fluorescence ± SD | Mean ratio |
|---|---|---|---|
| Mtb *inhA* WT | 2.723 ± 0.176 | 0.208 ± 0.043 | 13.69 ± 3.08 |
| Mtb *inhA* MUT | 2.462 ± 0.150 | 0.579 ± 0.082 | 4.38 ± 0.92 |
| Mtb *katG* WT | 1.621 ± 0.344 | 0.535 ± 0.161 | 3.15 ± 0.39 |
| Mtb *katG* MUT | 2.523 ± 0.161 | 0.664 ± 0.162 | 3.99 ± 0.77 |

**Supplementary Table S5. Allele discrimination of Mtb *inhA* and *katG* crRNAs *in vitro*.** On-target and off-target activities represent mean normalized fluorescence ± SD at 60 min. Activity ratios were calculated for each biological replicate as on-target/off-target fluorescence and are reported as mean ± SD. Data represents three independent biological replicates.

| Sub-panel | Target | n | Pearson *r* (95% CI) | *P* |
|---|---|---|---|---|
| **Malaria** | *P. falciparum* | 5 | −0.86 (−0.99 to 0.09) | 0.062 |
| | *P. vivax* | 5 | −0.77 (−0.98 to 0.35) | 0.126 |
| | *P. ovale* | 4 | −0.80 (−1.00 to 0.70) | 0.202 |
| | *P. malariae* | 5 | −0.75 (−0.98 to 0.39) | 0.145 |
| **HIV** | HIV-1 | 5 | −0.88 (−0.99 to 0.02) | 0.050 |
| | HIV-2 | 5 | −0.88 (−0.99 to 0.01) | 0.049 |
| **Tuberculosis** | Mtb IS6110 | 5 | −0.78 (−0.98 to 0.34) | 0.123 |
| | Mtb *inhA* WT | 4 | −0.89 (−1.00 to 0.51) | 0.115 |
| | Mtb *katG* WT | 5 | −0.93 (−1.00 to −0.30) | 0.020 |
| | Mtb *inhA* MUT | 5 | −0.92 (−0.99 to −0.18) | 0.029 |
| | Mtb *katG* MUT | 5 | −0.96 (−1.00 to −0.55) | 0.008 |

**Supplementary Table S6. Correlation between CARMEN fluorescence and qPCR Ct across HTM assays.** Pearson correlations were calculated using paired CARMEN fluorescence and determined qPCR Ct values (Ct < 38). qPCR-undetermined measurements were excluded.

| Matrix | Target | On-target crRNA mean normalized fluorescence ± SD | Off-target crRNA mean normalized fluorescence ± SD | Mean ratio ± SD |
|---|---|---|---|---|
| **Sputum** | *inhA* WT | 0.898 ± 0.050 | 0.099 ± 0.007 | 9.05 ± 0.87 |
| | *inhA* MUT | 0.782 ± 0.155 | 0.279 ± 0.018 | 2.80 ± 0.55 |
| | *katG* WT | 0.723 ± 0.053 | 0.124 ± 0.016 | 5.93 ± 0.99 |
| | *katG* MUT | 0.453 ± 0.136 | 0.187 ± 0.068 | 2.99 ± 2.11 |
| **Whole blood** | *inhA* WT | 0.806 ± 0.049 | 0.147 ± 0.004 | 5.49 ± 0.34 |
| | *inhA* MUT | 0.806 ± 0.079 | 0.230 ± 0.009 | 3.52 ± 0.44 |
| | *katG* WT | 0.704 ± 0.096 | 0.235 ± 0.021 | 2.99 ± 0.26 |
| | *katG* MUT | 1.300 ± 0.069 | 0.149 ± 0.023 | 8.92 ± 1.46 |
| **DBS** | *inhA* WT | 0.888 ± 0.064 | 0.148 ± 0.016 | 6.08 ± 0.77 |
| | *inhA* MUT | 0.762 ± 0.159 | 0.178 ± 0.009 | 4.30 ± 1.07 |
| | *katG* WT | 0.923 ± 0.098 | 0.211 ± 0.036 | 4.47 ± 0.86 |
| | *katG* MUT | 0.950 ± 0.104 | 0.232 ± 0.033 | 4.17 ± 0.76 |

**Supplementary Table S7. Allele discrimination of Mtb inhA and katG crRNAs across sample matrices.** On-target and off-target activities represent mean normalized fluorescence ± SD at 60 min. Activity ratios were calculated for each contrived sample as on-target/off-target fluorescence and are reported as mean ± SD. Data represents 20 contrived samples measured in technical duplicate per matrix.

| Target | Matrix | Input (copies µL$^{-1}$) | CARMEN positive | qPCR positive |
|---|---|---|---|---|
| ***P. falciparum*** | **Whole blood** | **$10^3$** | **18/20** | **15/20** |
| | | $10^2$ | 18/20 | 14/20 |
| | | $10^1$ | 18/20 | 7/20 |
| | | $10^0$ | 10/20 | 2/20 |
| | **Serum** | $10^3$ | 15/20 | 16/20 |
| | | $10^2$ | 14/20 | 14/20 |
| | | $10^1$ | 14/20 | 14/20 |
| | | $10^0$ | 15/20 | 4/20 |
| | **DBS** | $10^3$ | 19/20 | 11/20 |
| | | $10^2$ | 18/20 | 15/20 |
| | | $10^1$ | 17/20 | 8/20 |
| | | $10^0$ | 14/20 | 3/20 |
| **HIV-1** | **Whole blood** | **$10^3$** | **20/20** | **20/20** |
| | | $10^2$ | 19/20 | 18/20 |
| | | $10^1$ | 19/20 | 4/20 |
| | | $10^0$ | 19/20 | 0/20 |
| | **Serum** | $10^3$ | 18/20 | 12/20 |
| | | $10^2$ | 20/20 | 14/20 |
| | | $10^1$ | 16/20 | 14/20 |
| | | $10^0$ | 16/20 | 3/20 |
| | **DBS** | $10^3$ | 17/20 | 15/20 |
| | | $10^2$ | 19/20 | 12/20 |
| | | $10^1$ | 18/20 | 7/20 |
| | | $10^0$ | 16/20 | 4/20 |
| **Mtb IS6110** | **Sputum** | **$10^3$** | **20/20** | **20/20** |
| | | $10^2$ | 20/20 | 18/20 |
| | | $10^1$ | 20/20 | 12/20 |
| | | $10^0$ | 20/20 | 13/20 |
| | | $10^{-1}$ | 20/20 | 3/20 |
| | **Whole blood** | $10^3$ | 18/20 | 18/20 |
| | | $10^2$ | 18/20 | 16/20 |
| | | $10^1$ | 17/20 | 14/20 |
| | | $10^0$ | 17/20 | 13/20 |
| | | $10^{-1}$ | 15/20 | 4/20 |
| | **DBS** | $10^3$ | 19/20 | 16/20 |
| | | $10^2$ | 19/20 | 19/20 |
| | | $10^1$ | 17/20 | 15/20 |
| | | $10^0$ | 15/20 | 12/20 |
| | | $10^{-1}$ | 14/20 | 4/20 |

**Supplementary Table S8. Detection frequency of HTM CARMEN and qPCR across contrived sample matrices.** Values indicate the number of samples classified as positive out of 20 at each target concentration. CARMEN samples were classified as positive when normalized fluorescence exceeded the assay positivity threshold, defined as the mean no-template control fluorescence plus three standard deviations.

### Supporting Figures

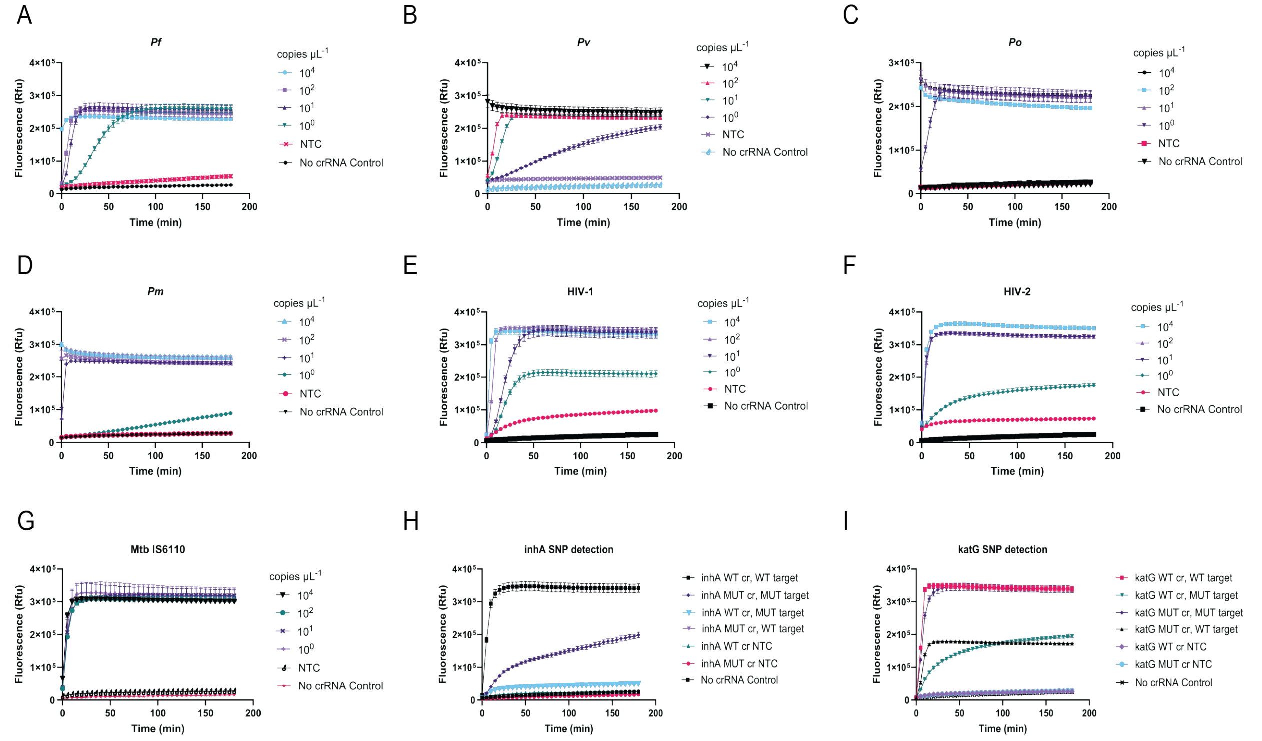

**Figure S1. HTM assay *in vitro* performance against synthetic DNA targets.** HTM crRNAs evaluated in singleplex SHERLOCK detection reactions against synthetic DNA of **(A)** *Plasmodium falciparum 18S,* **(B)** *Plasmodium vivax 18S,* **(C)** *Plasmodium ovale 18S,* **(D)** *Plasmodium malariae 18S,* **(E)** HIV-1, **(F)** HIV-2*,* **(G)** Mtb IS6110, **(H)** Mtb *inhA* ($10^4$ copies $\mu L^{-1}$)*, and* **(I)** Mtb *katG* ($10^4$ copies $\mu L^{-1}$). Each reaction was performed with serial dilutions of target input at $10^4$, $10^2$, $10^1$, and $10^0$ copies $\mu L^{-1}$, along with a No-Target Control (NTC) and a no crRNA control. Fluorescence signal (Rfu) was measured every 5 minutes over a period of 3 hours using Applied Biosystems Quantstudio 5; Filter Channel 1 (X1-M1) for FAM detection. Data are shown as mean ± SD and are representative of 3 biological replicates.

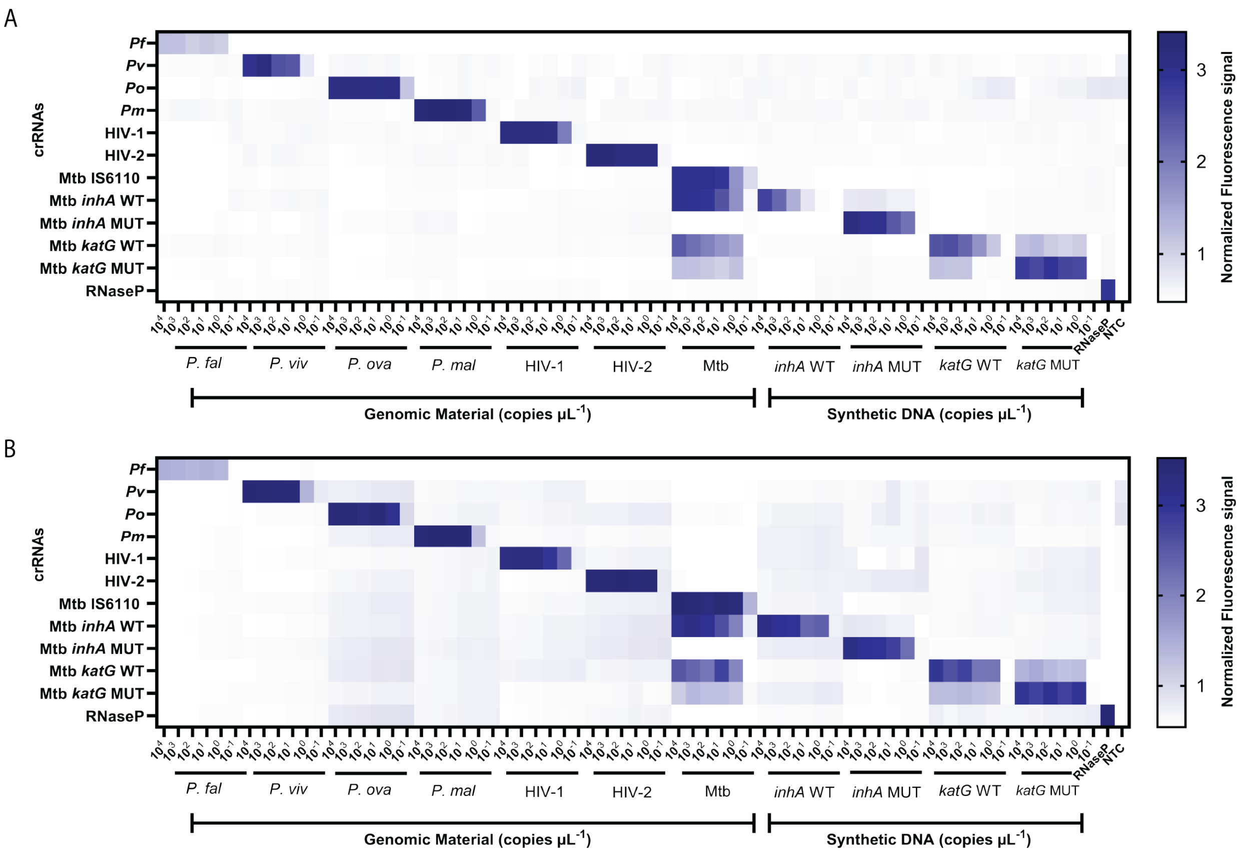

**Figure S2. *In vitro* analytical performance of the HTM CARMEN panel.** Sensitivity and specificity assessment of the HTM CARMEN assays under *in vitro* conditions. Heatmap shows normalized fluorescence for the indicated genomic or synthetic targets across concentrations ranging from $10^4$ to $10^{-1}$ copies $\mu L^{-1}$, displaying target-dependent signal generation of the corresponding Cas13a crRNAs at (**A**) 90 minutes or (**B**) 120 minutes post-reaction initiation. NTC = No-Target Control.

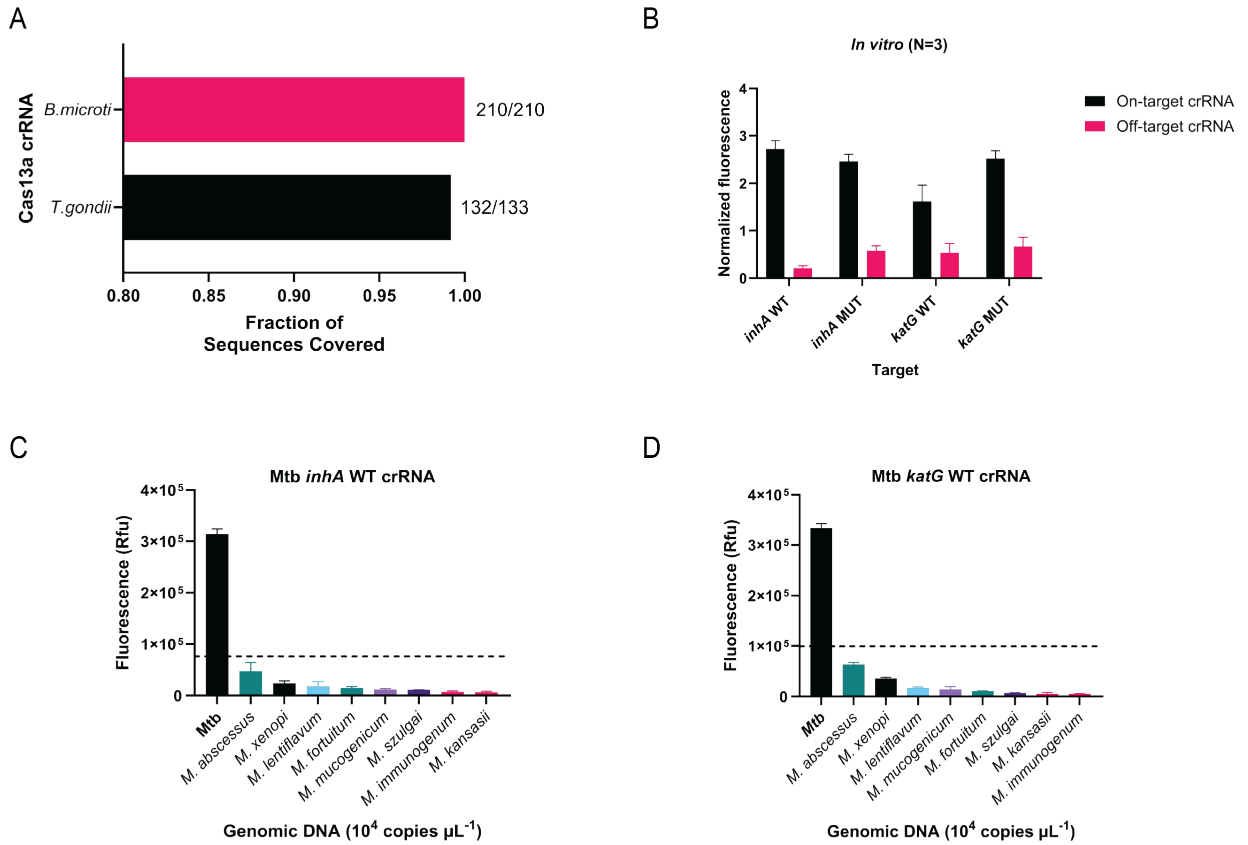

**Figure S3. Analytical specificity of Mtb resistance-associated crRNAs against nontuberculous mycobacteria. (A)** Predicted sequence coverage of crRNAs targeting *Toxoplasma gondii* and *Babesia microti*, as related apicomplexan specificity controls. **(B)** On-target and off-target fluorescence of allele-specific Mtb *inhA* and *katG* crRNAs at 60 min using wild-type Mtb genomic DNA or synthetic mutant gBlocks at $10^4$ copies $\mu L^{-1}$ under *in vitro* conditions. Data are shown as mean ± SD from three independent biological replicates. **(C-D)** Specificity of the Mtb (**C**) *inhA* WT and (**D**) *katG* WT crRNAs against Mtb and eight nontuberculous mycobacterial species (*M. abscessus, M. xenopi, M. lentiflavum, M. fortuitum, M. mucogenicum, M. szulgai, M. immunogenum,* and *M. kansasii*) at $10^4$ copies $\mu L^{-1}$. Dashed lines indicate the assay-specific positivity threshold, defined as the mean No-Target Control (NTC) fluorescence plus three standard deviations.

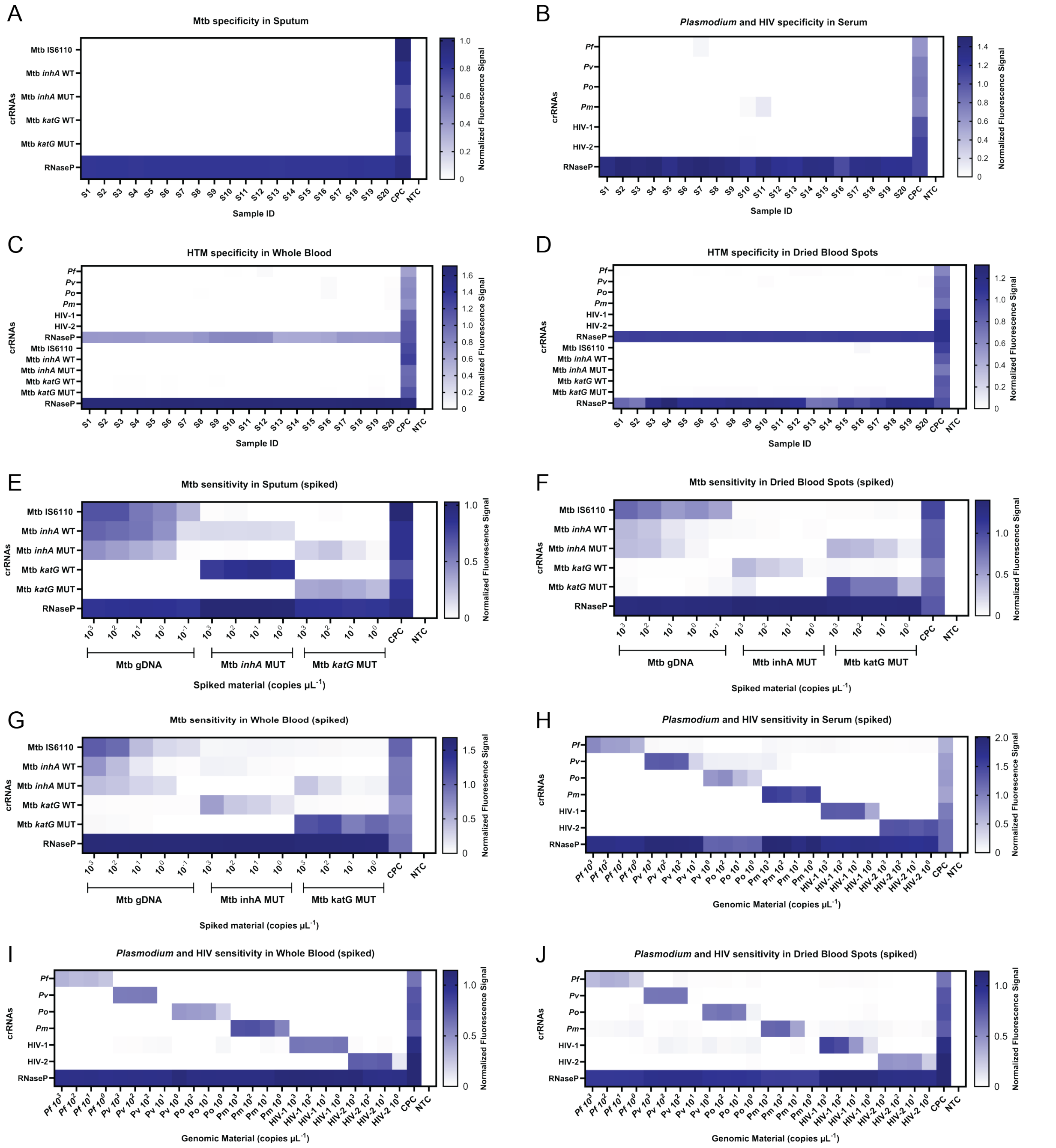

**Figure S4. Specificity of the HTM CARMEN panel across clinically relevant sample matrices.** (**A-D**) Normalized fluorescence in unspiked (**A**) sputum, (**B**) serum, (**C**) whole blood, and (**D**) dried blood spot (DBS) matrices tested against the indicated HTM crRNA panels. (**E-G**) Mtb genomic DNA and synthetic *inhA* MUT and *katG* MUT targets were spiked at $10^3$-$10^0$ copies $\mu L^{-1}$ into (**E**) sputum, (**F**) DBS, and (**G**) whole blood and tested against the Mtb crRNA panel. (**H-J**) HIV and *Plasmodium* genomic material were spiked at $10^3$-$10^0$ copies $\mu L^{-1}$ into (**H**) serum, (**I**) whole blood, and (**J**) DBS and tested against the HIV/*Plasmodium* crRNA panel. Heatmap values represent the mean of three technical replicates from one independent experiment. CPC, Combined Positive Control; NTC, No-Target Control.

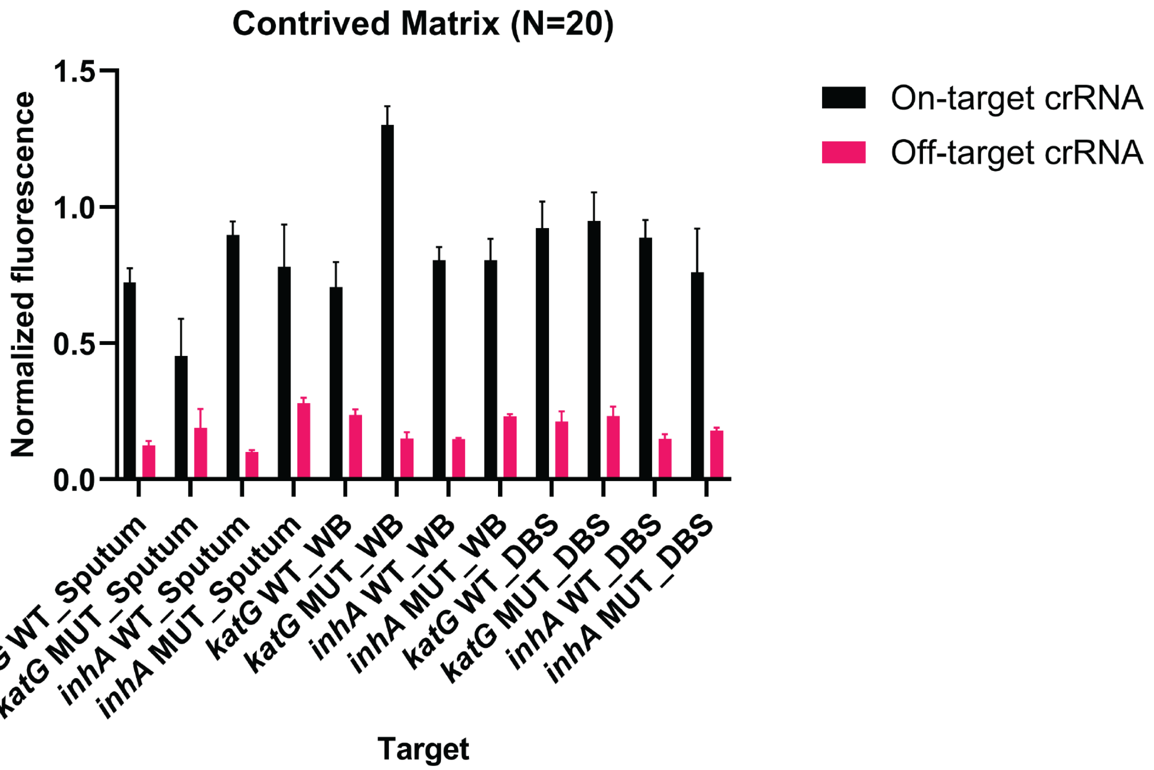

**Figure S5. Allele-specific fluorescence on HTM CARMEN across clinically relevant sample matrices.** On-target and off-target normalized fluorescence at 60 min for Mtb *inhA* WT, *inhA* MUT, *katG* WT, and *katG* MUT crRNAs in contrived sputum, whole blood, and dried blood spot (DBS) samples. Data are shown as mean ± SD across 20 contrived samples.

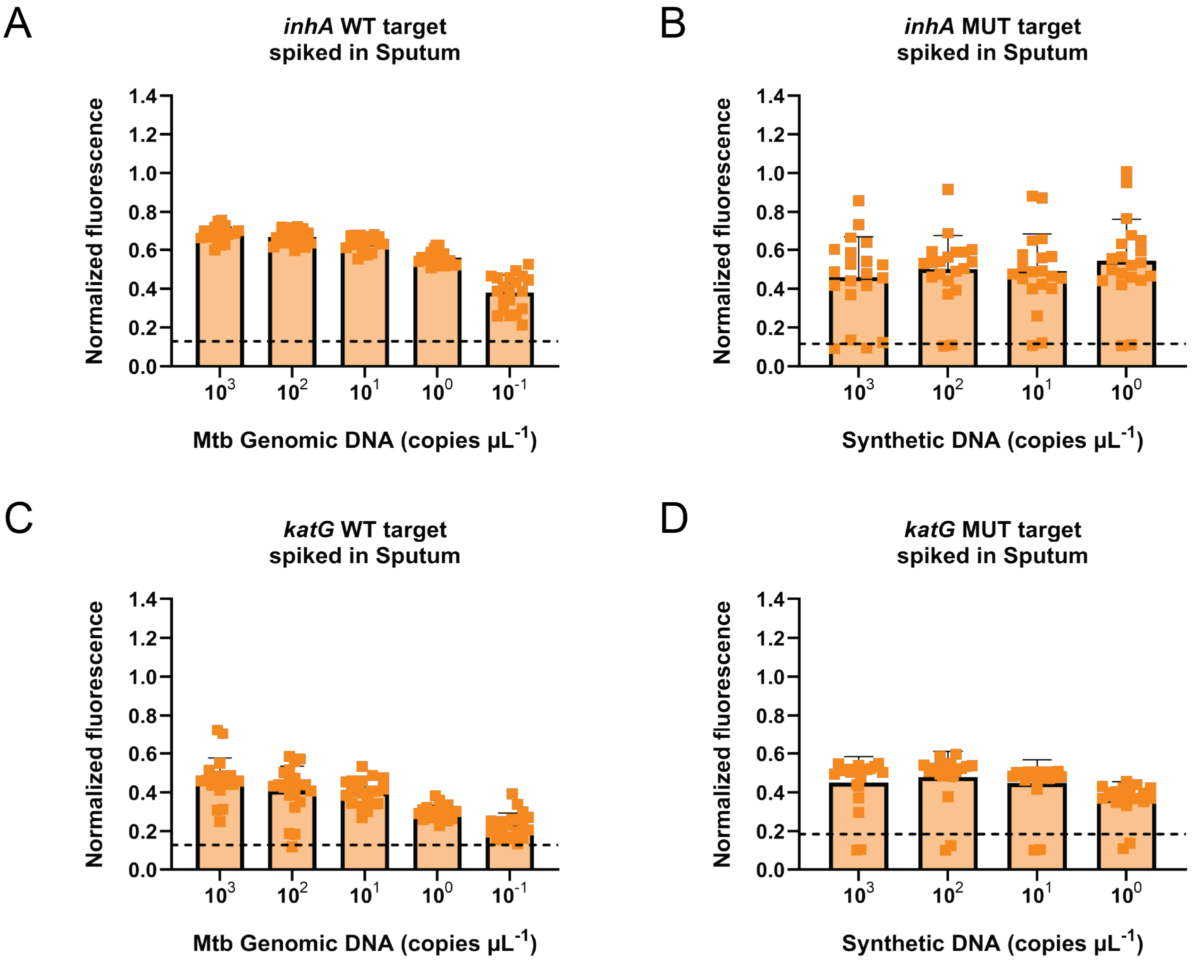

**Figure S6. Detection performance of the Mtb sub-panel in contrived sputum samples.** Normalized fluorescence values at 60 min for (**A**) *inhA* WT, (**B**) *inhA* MUT, (**C**) *katG* WT and (**D**) *katG* MUT targets at the indicated input concentrations. Twenty replicates were performed for each target in healthy human sputum samples. Bars show mean normalized CARMEN fluorescence ± SD; individual points represent each contrived sample. Dashed lines indicate assay-specific positivity thresholds.

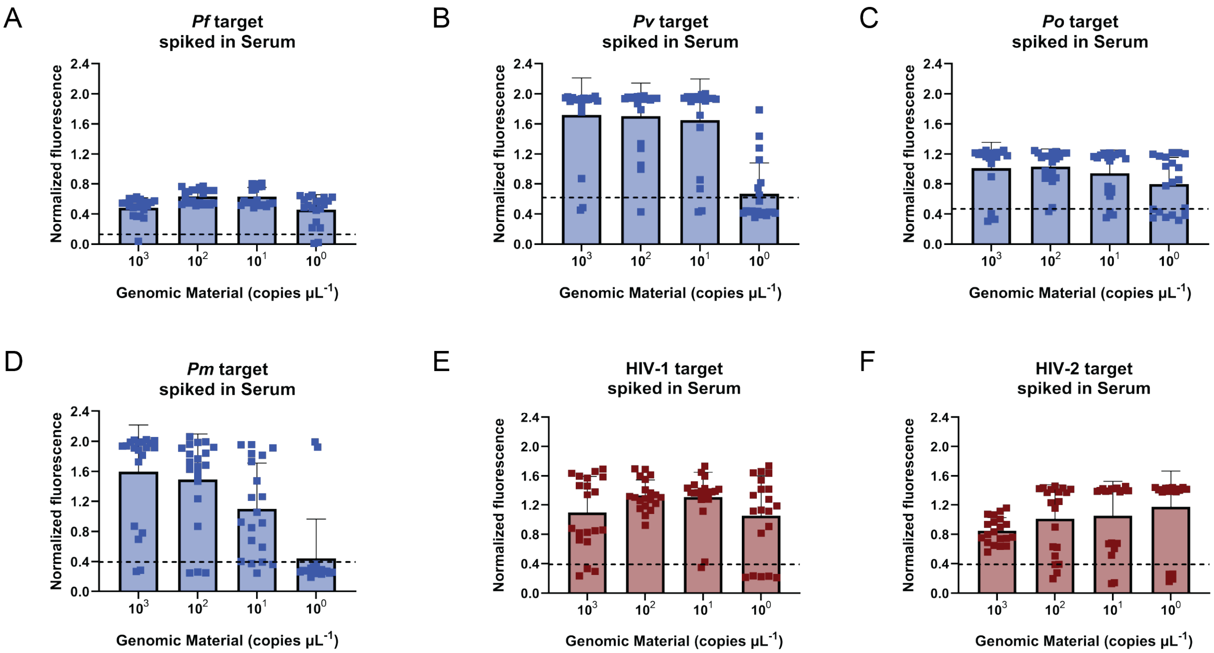

**Figure S7. Detection performance of the malaria and HIV sub-panels in contrived serum samples.** Normalized fluorescence values at 60 min for detection of low-abundance targets across 20 contrived samples containing (**A-D**) *P. falciparum*, *P. vivax*, *P. ovale*, and *P. malariae* genomic material, (**E-F**) HIV-1, and HIV-2 genomic RNA at the indicated input concentrations. Bars show mean normalized CARMEN fluorescence ± SD; individual points represent each contrived sample. Dashed lines indicate assay-specific positivity thresholds.

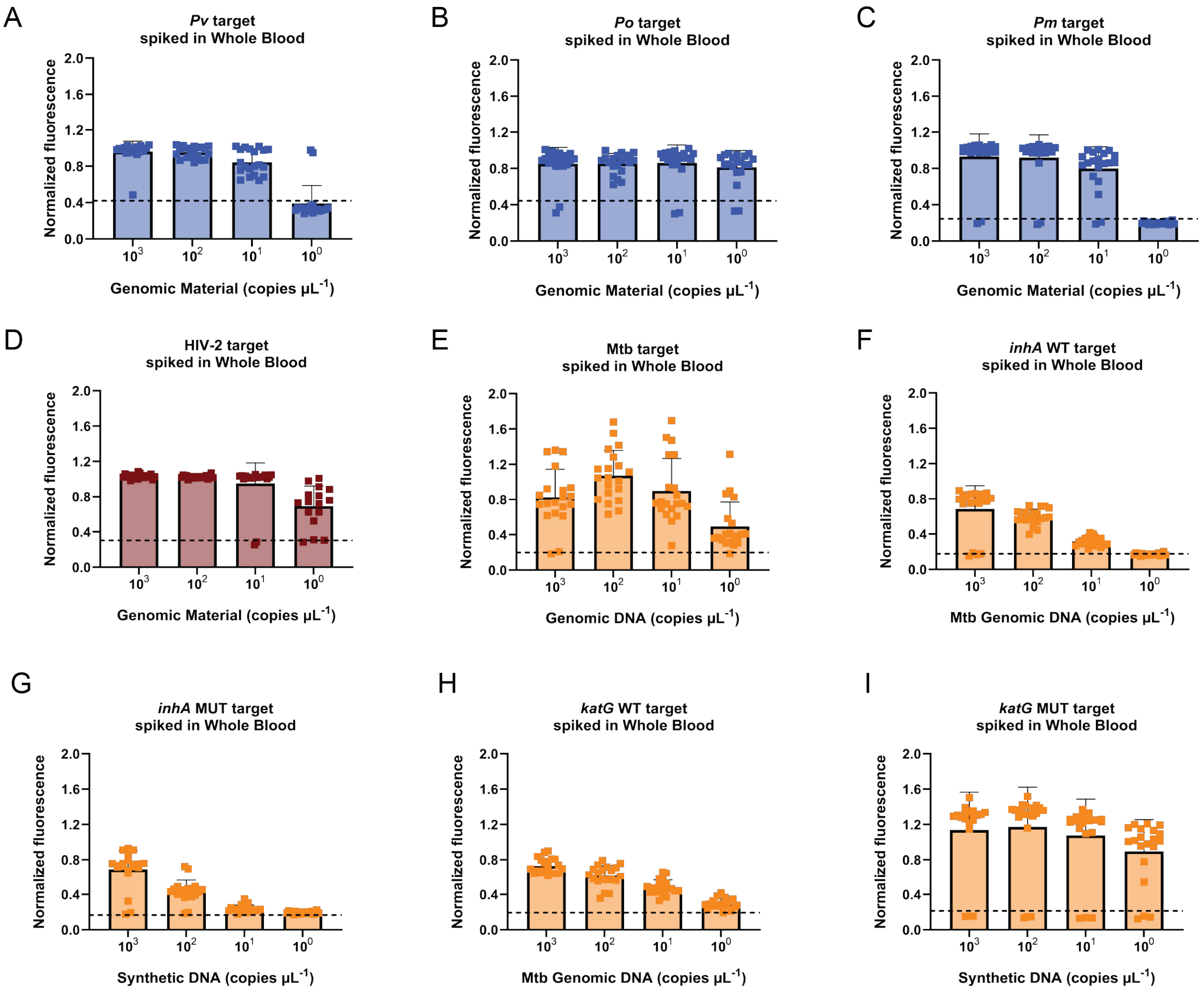

**Figure S8. Detection performance of HTM panel in contrived Whole Blood samples.** Normalized fluorescence values at 60 min for detection of low-abundance targets across 20 contrived samples containing (**A-C**) *Plasmodium vivax, P. ovale*, and *P. malariae* targets (**D**) HIV-2, and (**E-I**) Mtb panel targets at the indicated input concentrations. Bars show mean normalized CARMEN fluorescence ± SD; individual points represent each contrived sample. Dashed lines indicate assay-specific positivity thresholds.

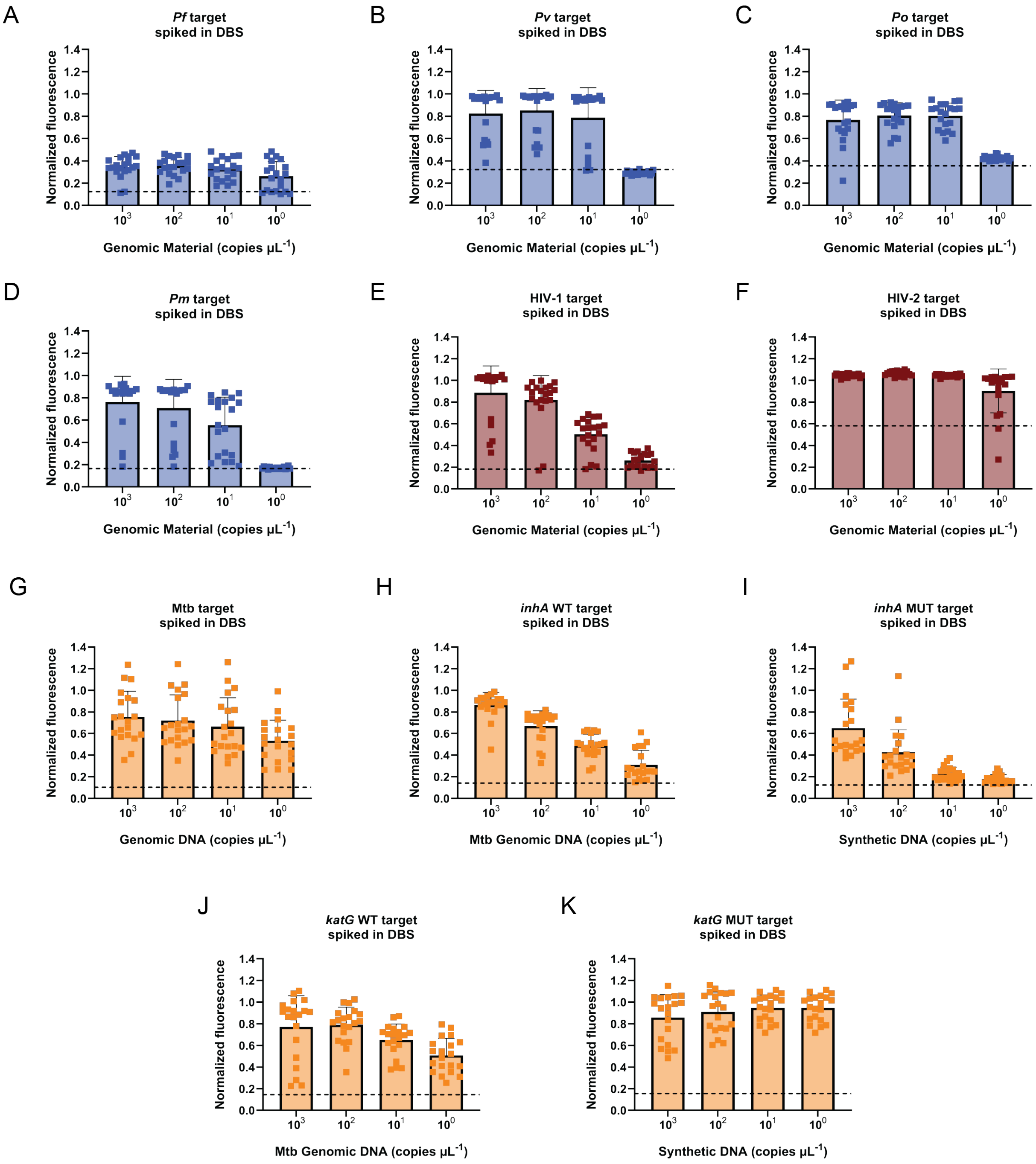

**Figure S9. Detection performance of HTM panel in contrived DBS samples.** Normalized fluorescence values at 60 min for detection of low-abundance targets across 20 contrived samples containing (**A-D**) *Plasmodium falciparum, P. vivax, P. ovale*, and *P. malariae* targets (**E-F**) HIV-1, HIV-2, and (**G-K**) Mtb panel targets at the indicated input concentrations. Bars show mean normalized CARMEN fluorescence ± SD; individual points represent each contrived sample. Dashed lines indicate assay-specific positivity thresholds.

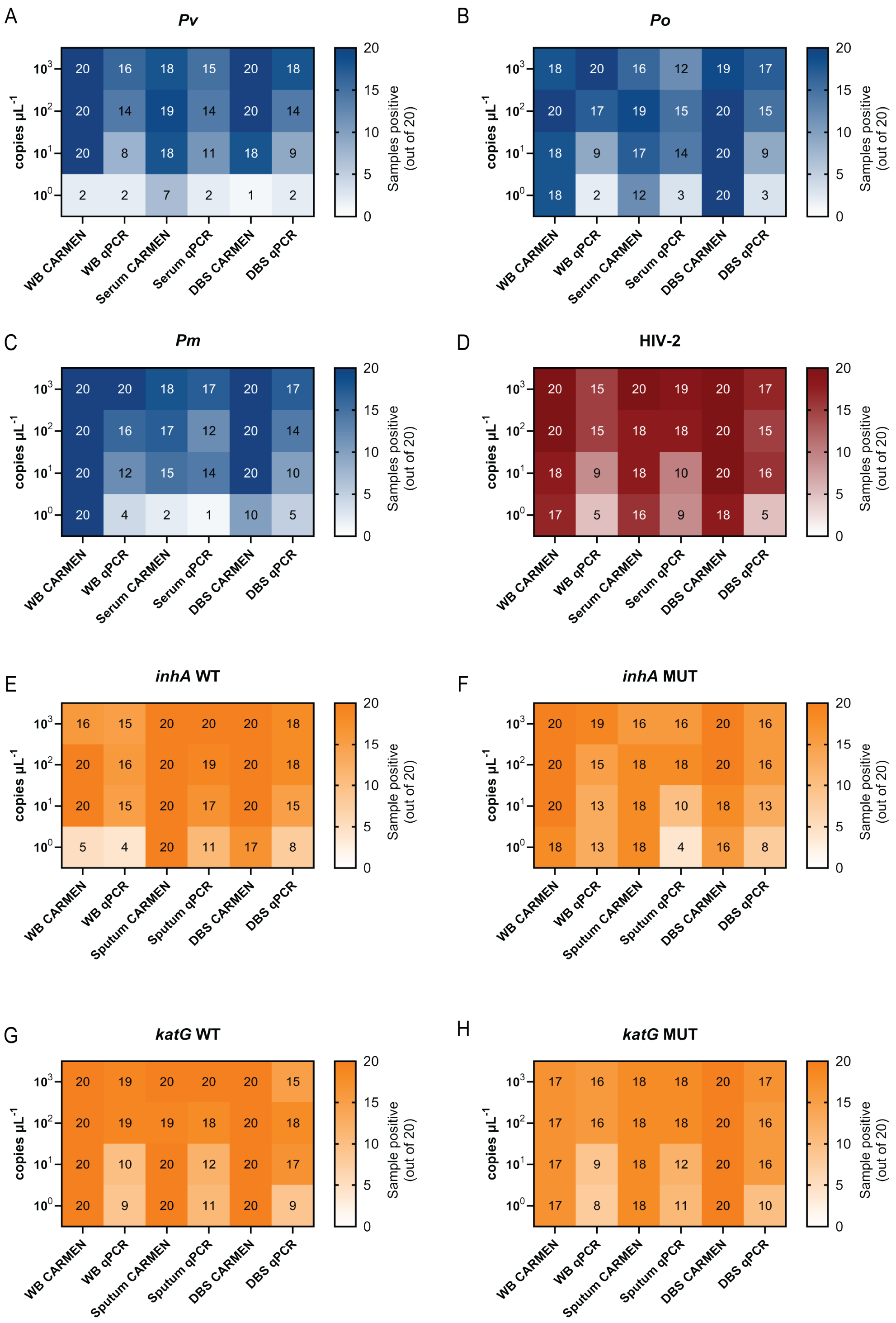

**Figure S10. Matrix comparison grids for CARMEN and qPCR.** Comparison of CARMEN and qPCR detection across target concentration and sample matrix for malaria sub-panel **(A)** *P. vivax*, **(B)** *P. ovale*, **(C)** *P. malariae*, HIV sub-panel **(D)** HIV-2*,* and Mtb sub-panel **(E)** *inhA* WT, **(F)** *inhA* MUT, **(G)** *katG* WT, **(H)** *katG* MUT. Heatmaps show the number of samples classified as positive out of 20 at each target concentration in whole blood, serum, and DBS for malaria and HIV sub-panels, sputum, whole blood, and DBS for Mtb sub-panel. Color scales reflect the number of samples classified as positive out of 20.

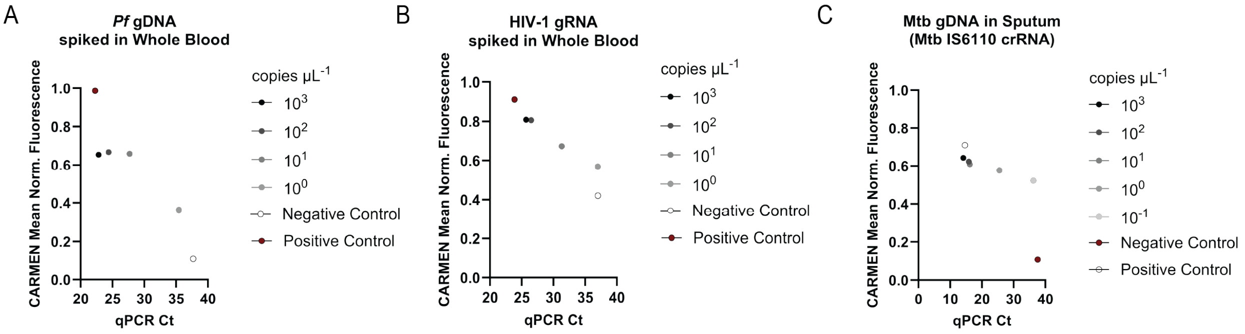

**Figure S11. Relationship between CARMEN fluorescence and qPCR Ct in contrived sample matrices. (A-C)** Relationship between CARMEN normalized fluorescence and qPCR cycle threshold (Ct) across target concentrations for **(A)** *P. falciparum* genomic DNA in whole blood, **(B)** HIV-1 genomic RNA in whole blood, and **(C)** Mtb genomic DNA detected with the IS6110 crRNA in sputum. Positive and negative controls are indicated.

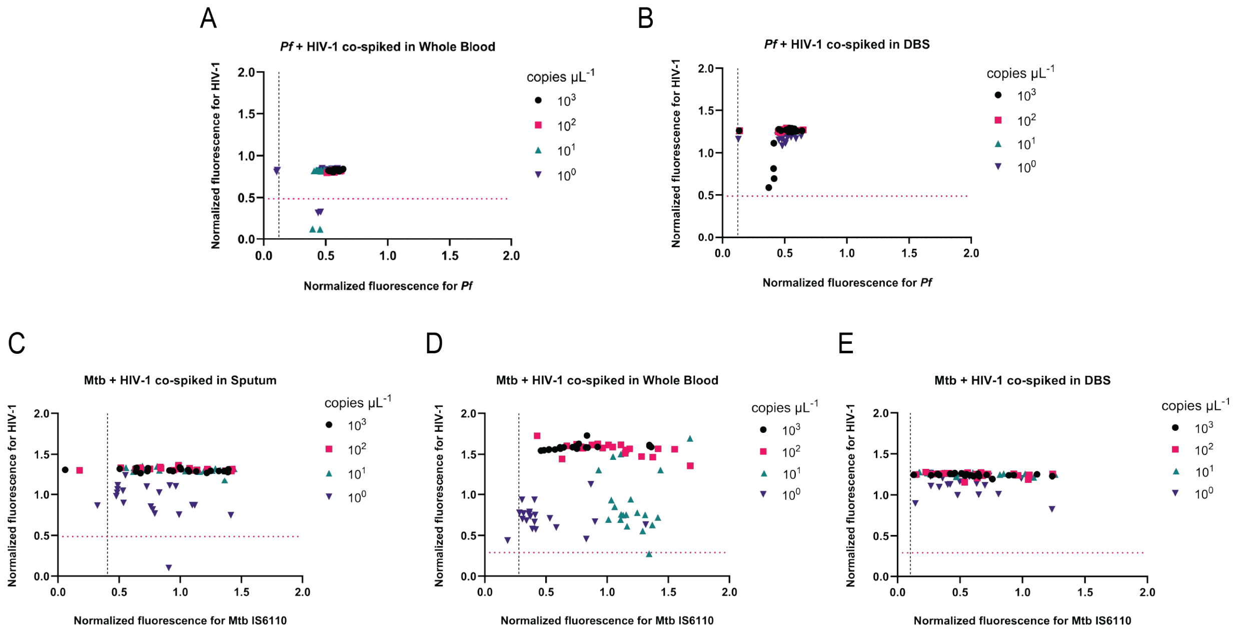

**Figure S12. Multiplexed detection of contrived *Pf*-HIV-1 and Mtb-HIV-1 coinfections.** Normalized fluorescence values at 60 min for detection of low-abundance targets across 20 contrived samples containing *Pf* and HIV-1 spiked simultaneously in **(A)** whole blood, **(B)** DBS, **(C-E)** Mtb and HIV-1 spiked simultaneously in **(C)** sputum, **(D)** whole blood, and **(E)** DBS, and at the indicated target concentrations. Each data point shows normalized CARMEN fluorescence for *Pf*-HIV-1 and Mtb-HIV-1 assays respectively; individual points represent each contrived sample. Dashed lines indicate assay-specific positivity thresholds for each respective assay.

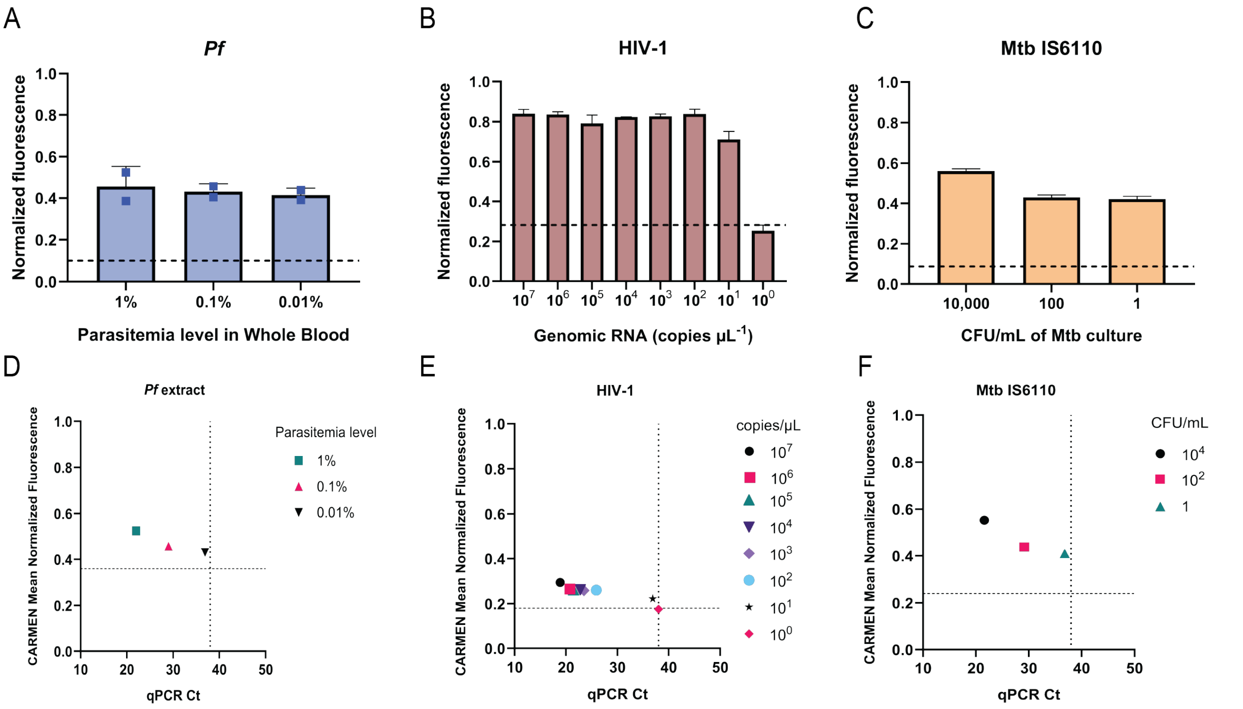

**Figure S13. Detection of extracted genomic material from bacterial, parasitic, and viral cultures across infection levels.** Detection of **(A)** *Plasmodium falciparum* genomic RNA extracted from whole blood cultures at decreasing parasitemia levels of 1%, 0.1%, and 0.01%. **(B)** Detection of HIV-1 viral RNA across a range of input concentrations ranging from $10^7$-$10^0$ copies μL$^{-1}$. **(C)** Detection of *Mycobacterium tuberculosis* using IS6110 targeting from bacterial cultures spanning 10,000 to 1 CFU mL$^{-1}$. (**D-F)** Concordance between CARMEN and qPCR for the detection of **(D)** *Plasmodium falciparum* genomic RNA extracted from whole blood cultures at decreasing parasitemia levels of 1%, 0.1%, and 0.01%. (**E)** HIV-1 viral RNA extracted from viral cultures ranging from $10^7$ to $10^0$ copies μL$^{-1}$. **(F)** *Mycobacterium tuberculosis* targeting IS6110 from bacterial cultures spanning 10,000 to 1 CFU mL$^{-1}$. Fluorescence signals are shown as normalized ratios relative to no-template controls.

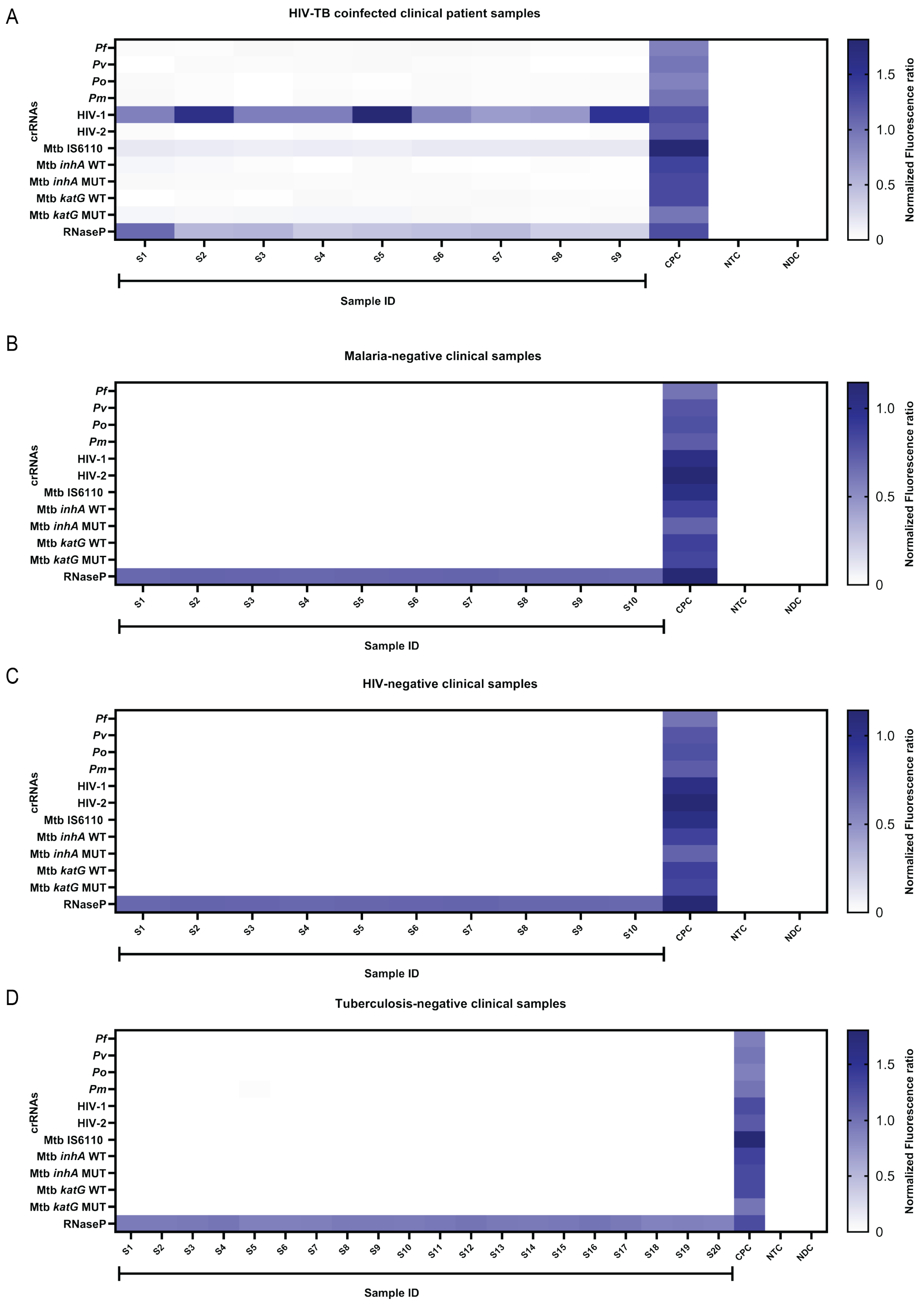

**Figure S14. Detection of malaria, HIV-1, TB, and HIV-TB coinfection in clinical specimens. (A)** Analysis of 9 confirmed-positive HIV-TB coinfected clinical specimens showing normalized CARMEN fluorescence across the indicated HTM crRNAs. Specificity testing of confirmed-negative specimens showing normalized CARMEN fluorescence across the indicated HTM crRNAs for (B) 10 malaria-negative whole blood specimens, (C) 10 HIV-negative whole blood specimens, and (D) 20 Mtb-negative specimens. Negative normalized fluorescence values reflect signal below the background reference used for normalization. CPC = Combined Positive Control. NTC = No-Target Control. NDC = No Detection Control.

### Supplemental Methods

#### M1. Protocol for Nucleic acid extraction using Quick DNA/RNA™ MagBead Kit

- **Pre-treatment of Patient Samples:**

1. Add 200µL of 2X Concentrate DNA/RNA Shield™ per 100µL of patient sample and mix well.
2. Add 8µL of Proteinase K to each sample and mix well. Incubate at 30°C for 30 minutes.
3. Vortex samples and centrifuge at maximum speed (16,000 x g) for 2 minutes to pellet debris.
4. Transfer the clear supernatant into a new nuclease-free tube and discard any precipitate.
5. Add 400µL of 100% isopropanol and mix well.
6. Store the product on ice until use and continue to the "Total Nucleic Acid Purification" section of Zymo Research Quick-DNA/RNA™ MagBead protocol.

- **Total Nucleic Acid purification:**

1. Add 200 µL DNA/RNA Lysis Buffer to 200 µL sample and mix well.
2. Add 400 µL ethanol (95-100%) to the sample (1:1) and mix well.
3. Add 30 µL MagBinding Beads to the sample and mix well for 30-40 minutes on a plate rotator.
4. Transfer the plate to the magnetic stand until beads have pelleted and then aspirate and discard the cleared supernatant.
5. Add 500 µL MagBead DNA/RNA Wash 1 and mix well. Pellet the beads and discard the supernatant.
6. Add 500 µL MagBead DNA/RNA Wash 2 and mix well. Pellet the beads and discard the supernatant.
7. Add 500 µL ethanol (95-100%) and mix well. Pellet the beads and discard the supernatant. Repeat this step.
8. Dry the beads for 30 minutes or until dry.
9. To elute DNA/RNA from the beads, add 50 µL DNase/RNase-Free Water and mix well. Incubate for 5-10 minutes.
10. Transfer the plate to the magnetic stand until beads have pelleted, then aspirate and dispense the eluted DNA/RNA to a new Elution Plate/tube.
11. Post extraction, properly dispose of all plates except the Elution Plate. Store Elution Plate on ice or at -80°C until proceeding to amplification steps.

#### *M2.* *In vitro* transcription of crRNAs

- Synthetic DNA oligonucleotides corresponding to HTM panel crRNAs were procured from Thermo Fisher Scientific and used as templates for *in vitro* transcription (IVT) with the HiScribe T7 High Yield RNA Synthesis Kit (New England Biolabs) to generate crRNAs. A T7 promoter-containing primer was annealed to each DNA template before transcription. Reagents were then combined in the following volumes:

| **Reagent** | **Volume (µL)** |
|---|---|
| Nuclease Free Water | 5.46 |
| T7 Primer (100 µM) | 1 |
| 10x NEB Standard Taq Buffer | 1 |
| crRNA template (1 ug $\mu L^{-1}$) | 2.54 |

The above mixture was denatured at 95°C for 5 minutes and then slowly cooled (0.1°C $s^{-1}$) to 4°C. The following was then added to the above mixture:

| **Reagent** | **Volume (µL)** |
|---|---|
| NTP Buffer mix | 10 |
| T7 RNA Polymerase mix | 2 |
| Nuclease Free Water | 17 |

The 39 µL IVT reactions were incubated for 16 h at 37°C, followed by treatment with DNase I (New England Biolabs) to remove the DNA templates. Transcribed RNA products were purified using RNAClean XP beads (Beckman Coulter). Beads were added to the IVT reactions at a 10:3 ratio and supplemented with isopropanol at a 3:1 ratio relative to the IVT reaction volume. Purified RNA was quantified using the Qubit RNA Broad Range (BR) Assay Kit (Invitrogen) according to the manufacturer's instructions. Prior to use in detection assays, crRNAs were diluted to the required working concentrations.

#### M3. Preparation of Primer Pools

- Upon receipt, lyophilized primers were resuspended to a concentration of 100 µM in nuclease-free (non-DEPC-treated) water and stored at −20°C. Working primer solutions were prepared by diluting each stock to 5 µM. The 5 µM forward and reverse primers were then combined into two separate primer pools according to the ratios specified in the table below. Primer pools were aliquoted and stored at −20°C until use.

| **Primer** | **Ratio** | **Volume (µL)** |
|---|---|---|
| *Pf* | 3 | 450 |
| *Pv, Po, Pm* | 3 | 450 |
| HIV-1 | 2 | 300 |

| | | |
|---|---|---|
| HIV-2 | 2 | 300 |
| Mtb IS6110 | 3 | 450 |
| *inhA* | 2 | 300 |
| *katG* | 2 | 300 |
| RNaseP | 2 | 300 |
| **Total** | 19 | 2850 |
